# Opposite white matter abnormalities in post-infectious vs. gradual onset chronic fatigue syndrome revealed by diffusion MRI

**DOI:** 10.1101/2024.08.04.24311483

**Authors:** Qiang Yu, Richard A. Kwiatek, Peter Del Fante, Anya Bonner, Vince D. Calhoun, Grant A. Bateman, Takashi Yamamura, Zack Y. Shan

## Abstract

Myalgic encephalomyelitis/chronic fatigue syndrome (ME/CFS) is a complex and debilitating illness with an unknown pathogenesis. Although post-infectious (PI-ME/CFS) and gradual onset ME/CFS (GO-ME/CFS) manifest similar symptoms, it has long been suspected that different disease processes underlie them. However, the lack of biological evidence has left this question unanswered. In this study, we recruited PI-ME/CFS and GO-ME/CFS patients based on consensus diagnoses made by two experienced clinicians and compared their diffusion MRI features with those of rigorously matched healthy controls (HCs) with sedentary lifestyles. PI-ME/CFS patients showed significantly higher axial diffusivities (ADs) in several association and projection fibres compared to HCs. Higher AD values in PI-ME/CFS were significantly related to worse physical summary scores. In contrast, GO-ME/CFS patients exhibited significantly decreased ADs in the corpus callosum. Lower AD values in GO- ME/CFS patients were significantly associated with lower mental summary scores in commissural and projection fibres. Distinct patterns of AD alterations in PI-ME/CFS and GO- ME/CFS provide neurophysiological evidence of different disease processes and highlight the heterogeneities of ME/CFS. These results also help explain inconsistent findings in previous ME/CFS studies and guide future intervention design.

## Introduction

Myalgic encephalomyelitis/chronic fatigue syndrome (ME/CFS) is a complex, debilitating disorder characterised by persistent fatigue, cognitive impairment, and various other symptoms that significantly impact patients’ quality of life^1–5^. Despite its significant burden, the underlying pathophysiological mechanism of ME/CFS remains unclear, impeding the development of accurate diagnostic methods and effective treatments.

Two types of onset have been observed for ME/CFS^1,6^: post-infectious (PI-ME/CFS) and gradual onset (GO-ME/CFS). In PI-ME/CFS, symptoms develop rapidly following a viral or bacterial infection^7^, which can include Epstein-Barr virus, Ross River virus, Coxiella burnetii (Q fever), influenza, and other coronaviruses, with ME/CFS symptoms manifest after an infectious episode (64-80%)^6,8^. In contrast, GO-ME/CFS is characterised by a slow, progressive development of symptoms over months or even years, without a clear infectious trigger^1,6^. It has long been suspected that patients with these different onset types may have distinct pathophysiological mechanisms^1,6^. Nevertheless, this question has yet to be answered conclusively in the absence of biological evidence.

In recent years, advanced neuroimaging techniques, such as diffusion tensor imaging (DTI), have been used as powerful tools for investigating the structural and functional integrity of the brain across different neurological conditions^9^. Fractional anisotropy (FA), mean diffusivity (MD), axial diffusivity (AD), and radial diffusivity (RD) are commonly used DTI metrics that provide valuable information regarding the integrity of white matter (WM) in the brain. Alterations in these diffusion metrics have been observed in various neurological and psychiatric disorders, offering valuable insights into the underlying pathophysiological mechanisms^10–14^. However, previous DTI results for ME/CFS are inconsistent or even contradictory^15–19^, possibly due to bias from small sample sizes or the heterogeneity of ME/CFS. In this study, we investigated the WM in ME/CFS using a rigorous and systematic approach, following recommendations for high reliability in translational research^20^. By pre-registering study protocol^21^, recruiting PI-ME/CFS and GO-ME/CFS patients based on consensus diagnoses made by two experienced clinicians (RK and PDF)^21^, using rigorously matched healthy controls (HCs) with sedentary lifestyles^22^, and employing a transparent methodological approach, this study aimed to provide a more reliable and comprehensive understanding of the WM microstructural abnormalities associated with different onset types of ME/CFS using DTI. To the best of our knowledge, this is the first study to investigate WM microstructural abnormalities using DTI in different ME/CFS subgroups based on the onset of their illness. By characterising differences in DTI metrics between patients and controls and their associations with clinical symptoms, we aim to elucidate the potential neurobiological mechanisms underlying ME/CFS and guide future intervention design.

## Results

### Participant characteristics

Here, we analysed DTI data from 143 participants, including 76 patients with ME/CFS (mean age, 42.64 ± 12.71 [standard deviation]; 64 women) and 67 HCs (mean age, 37.51 ± 11.57 [standard deviation]; 52 women). PI-ME/CFS and GO-ME/CFS were determined by consensus of interviews independently performed by two experienced clinicians. The HCs were selected to match 1:1 in sample size, age, sex, and activity levels with each patient group. The PI- ME/CFS group consisted of 43 PI-ME/CFS participants (mean age, 42.58 ± 12.41 [standard deviation]; 37 women) and 43 HCs (mean age, 40.19 ± 8.29 [standard deviation]; 37 women) (Fig. 1). The GO-ME/CFS group consisted of 33 GO-ME/CFS participants (mean age, 42.03 ± 12.87 [standard deviation]; 26 women) and 33 HCs (mean age, 42.18 ± 7.35 [standard deviation]; 26 women). Although excluding additional eligible HCs might seem counterproductive, this matching minimises sampling bias and helps make clear and unambiguous conclusions about the impact of ME/CFS on DTI metrics. Furthermore, the results for all eligible participants for PI-ME/CFS (43 PI-ME/CFS and 58 HCs) and GO- ME/CFS (33 GO-ME/CFS and 66 HCs) groups before number matching (1:1) are provided in the supplementary information.

**Fig. 1:**
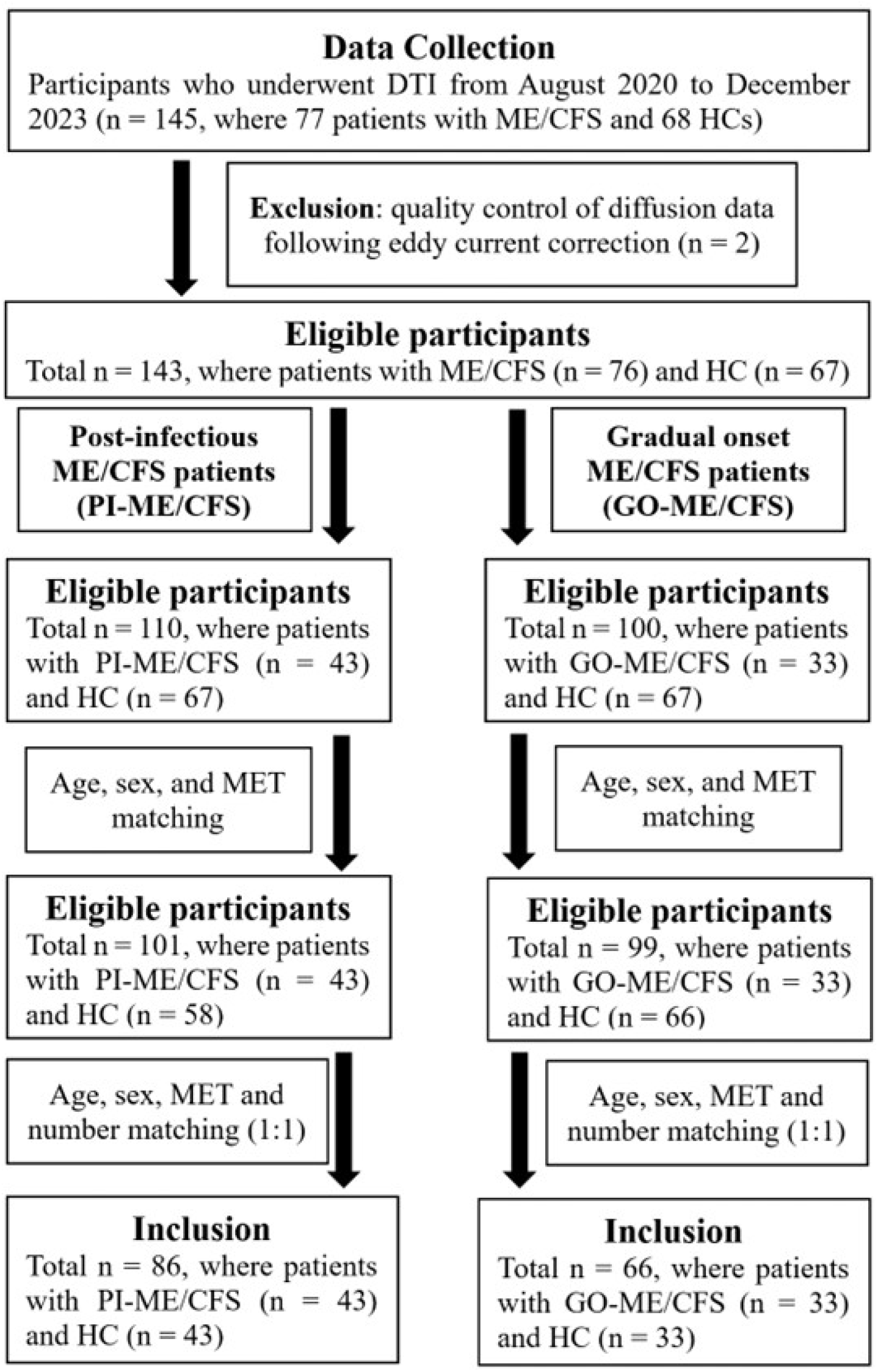
Flowchart of participants included in the study. A total of 86 participants who underwent diffusion tensor images (DTI) were included in the final analysis for post-infectious ME/CFS (PI-ME/CFS) group, and total of 66 for those gradual onset ME/CFS (GO-ME/CFS) group.

Table 1 presents the demographic characteristics and behavioural assessments of the PI- ME/CFS group. There are no significant differences in age (P = 0.67), body mass index (BMI) (P = 0.82), or MET rate (P = 0.10) between the patients and HCs. Compared to HCs, the PI- ME/CFS patients have higher Hospital Anxiety and Depression Scale (HADS) anxiety (P < 0.001) and depression (P < 0.001) scores and lower 36-item Short-Form (SF-36) mental (P < 0.001) and physical (P < 0.001) component scores. The HCs also have better overall sleep quality (P < 0.001) and lower levels of disability (P < 0.001) than the patients.

**Table 1:**
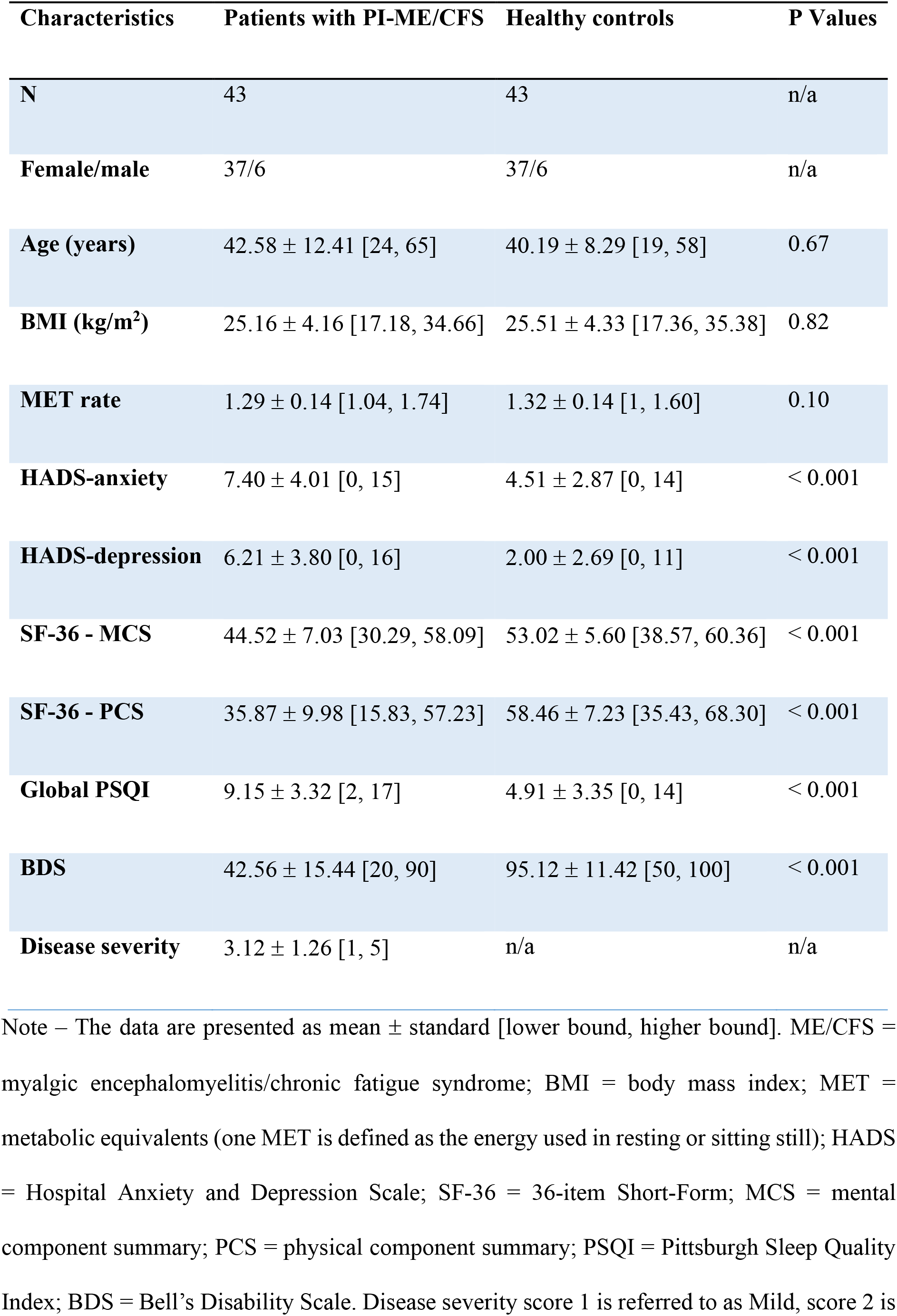
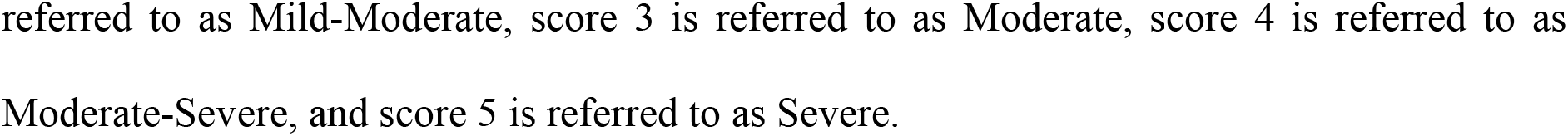
Demographic and behavioural information of the study participants in the final analysis for post-infectious ME/CFS (PI-ME/CFS) group.

| Characteristics | Patients with PI-ME/CFS | Healthy controls | P Values |
| --- | --- | --- | --- |
| **N** | 43 | 43 | n/a |
| **Female/male** | 37/6 | 37/6 | n/a |
| **Age (years)** | 42.58 ± 12.41 [24, 65] | 40.19 ± 8.29 [19, 58] | 0.67 |
| **BMI (kg/m<sup>2</sup>)** | 25.16 ± 4.16 [17.18, 34.66] | 25.51 ± 4.33 [17.36, 35.38] | 0.82 |
| **MET rate** | 1.29 ± 0.14 [1.04, 1.74] | 1.32 ± 0.14 [1, 1.60] | 0.10 |
| **HADS-anxiety** | 7.40 ± 4.01 [0, 15] | 4.51 ± 2.87 [0, 14] | < 0.001 |
| **HADS-depression** | 6.21 ± 3.80 [0, 16] | 2.00 ± 2.69 [0, 11] | < 0.001 |
| **SF-36 - MCS** | 44.52 ± 7.03 [30.29, 58.09] | 53.02 ± 5.60 [38.57, 60.36] | < 0.001 |
| **SF-36 - PCS** | 35.87 ± 9.98 [15.83, 57.23] | 58.46 ± 7.23 [35.43, 68.30] | < 0.001 |
| **Global PSQI** | 9.15 ± 3.32 [2, 17] | 4.91 ± 3.35 [0, 14] | < 0.001 |
| **BDS** | 42.56 ± 15.44 [20, 90] | 95.12 ± 11.42 [50, 100] | < 0.001 |
| **Disease severity** | 3.12 ± 1.26 [1, 5] | n/a | n/a |

Note – The data are presented as mean ± standard [lower bound, higher bound]. ME/CFS = myalgic encephalomyelitis/chronic fatigue syndrome; BMI = body mass index; MET = metabolic equivalents (one MET is defined as the energy used in resting or sitting still); HADS = Hospital Anxiety and Depression Scale; SF-36 = 36-item Short-Form; MCS = mental component summary; PCS = physical component summary; PSQI = Pittsburgh Sleep Quality Index; BDS = Bell's Disability Scale. Disease severity score 1 is referred to as Mild, score 2 is
referred to as Mild-Moderate, score 3 is referred to as Moderate, score 4 is referred to as Moderate-Severe, and score 5 is referred to as Severe.

The GO-ME/CFS group showed similar demographics and symptoms scores to the PI-ME/CFS group (Table 2). There are no significant differences in age (P = 0.75), BMI (P = 0.27), and MET rate (P = 0.09) between the GO-ME/CFS patients and HCs. GO-ME/CFS patients have higher HADS anxiety (P < 0.001) and depression (P < 0.001) scores and lower SF-36 mental (P < 0.001) and physical (P < 0.001) component scores. The HCs also have better overall sleep quality (P < 0.001) and lower levels of disability (P < 0.001) than the patients.

**Table 2:**
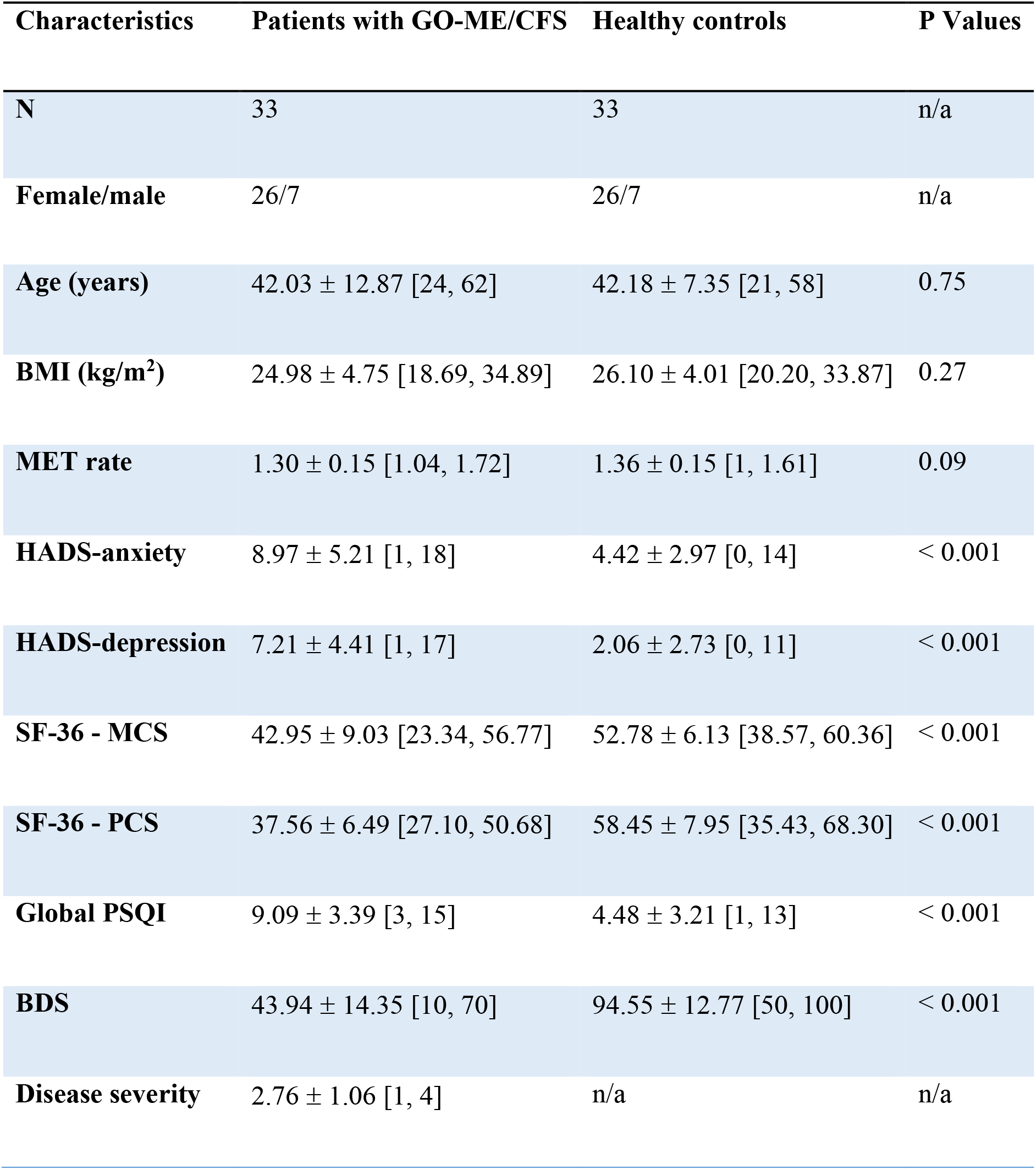
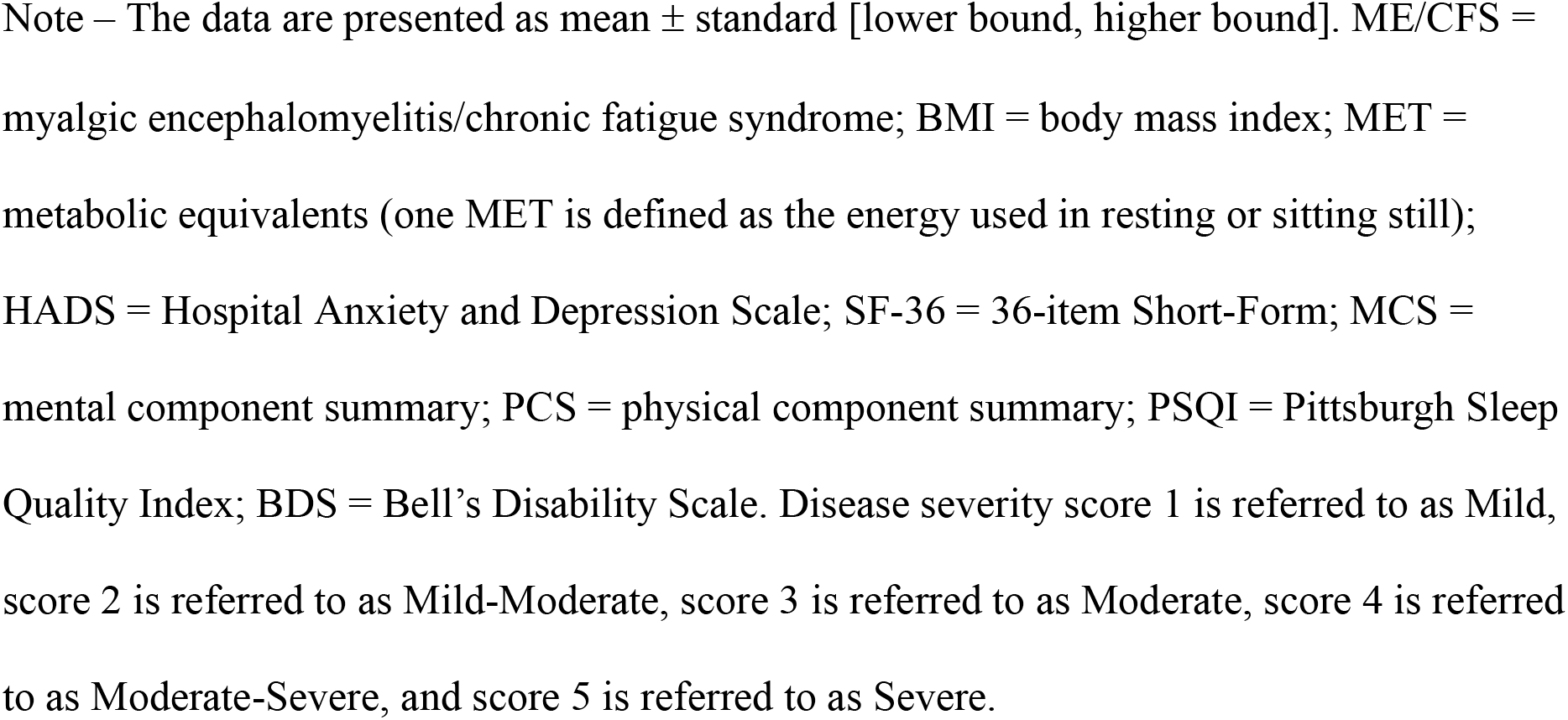
Demographic and behavioural information of the study participants in the final analysis for gradual onset ME/CFS (GO-ME/CFS) group.

| Characteristics | Patients with GO-ME/CFS | Healthy controls | P Values |
| --- | --- | --- | --- |
| **N** | 33 | 33 | n/a |
| **Female/male** | 26/7 | 26/7 | n/a |
| **Age (years)** | 42.03 ± 12.87 [24, 62] | 42.18 ± 7.35 [21, 58] | 0.75 |
| **BMI (kg/m<sup>2</sup>)** | 24.98 ± 4.75 [18.69, 34.89] | 26.10 ± 4.01 [20.20, 33.87] | 0.27 |
| **MET rate** | 1.30 ± 0.15 [1.04, 1.72] | 1.36 ± 0.15 [1, 1.61] | 0.09 |
| **HADS-anxiety** | 8.97 ± 5.21 [1, 18] | 4.42 ± 2.97 [0, 14] | < 0.001 |
| **HADS-depression** | 7.21 ± 4.41 [1, 17] | 2.06 ± 2.73 [0, 11] | < 0.001 |
| **SF-36 - MCS** | 42.95 ± 9.03 [23.34, 56.77] | 52.78 ± 6.13 [38.57, 60.36] | < 0.001 |
| **SF-36 - PCS** | 37.56 ± 6.49 [27.10, 50.68] | 58.45 ± 7.95 [35.43, 68.30] | < 0.001 |
| **Global PSQI** | 9.09 ± 3.39 [3, 15] | 4.48 ± 3.21 [1, 13] | < 0.001 |
| **BDS** | 43.94 ± 14.35 [10, 70] | 94.55 ± 12.77 [50, 100] | < 0.001 |
| **Disease severity** | 2.76 ± 1.06 [1, 4] | n/a | n/a |

Note – The data are presented as mean ± standard [lower bound, higher bound]. ME/CFS = myalgic encephalomyelitis/chronic fatigue syndrome; BMI = body mass index; MET = metabolic equivalents (one MET is defined as the energy used in resting or sitting still); HADS = Hospital Anxiety and Depression Scale; SF-36 = 36-item Short-Form; MCS = mental component summary; PCS = physical component summary; PSQI = Pittsburgh Sleep Quality Index; BDS = Bell's Disability Scale. Disease severity score 1 is referred to as Mild, score 2 is referred to as Mild-Moderate, score 3 is referred to as Moderate, score 4 is referred to as Moderate-Severe, and score 5 is referred to as Severe.

### Higher AD in PI-ME/CFS patients than HCs

Figure 2 illustrates the tract-based spatial statistics (TBSS) AD results between the HCs and PI-ME/CFS patient groups in Montreal Neurological Institute (MNI) 152 standard space. As shown in Fig. 2, the AD measures in the PI-ME/CFS patient group were significantly higher than those in the HCs in the following fibre tracts: association fibres (right superior longitudinal fasciculus, right uncinate fasciculus, and right external capsule), and projection fibres (right cerebellar peduncle, left inferior cerebellar peduncle, middle cerebellar peduncle, left superior cerebellar peduncle, right superior corona radiata, right posterior corona radiata, right sagittal stratum, corticospinal tract, and left medial lemniscus). There were no significant group differences in TBSS for FA, MD, and RD between patients and HCs in the PI-ME/CFS group.

**Fig. 2.**
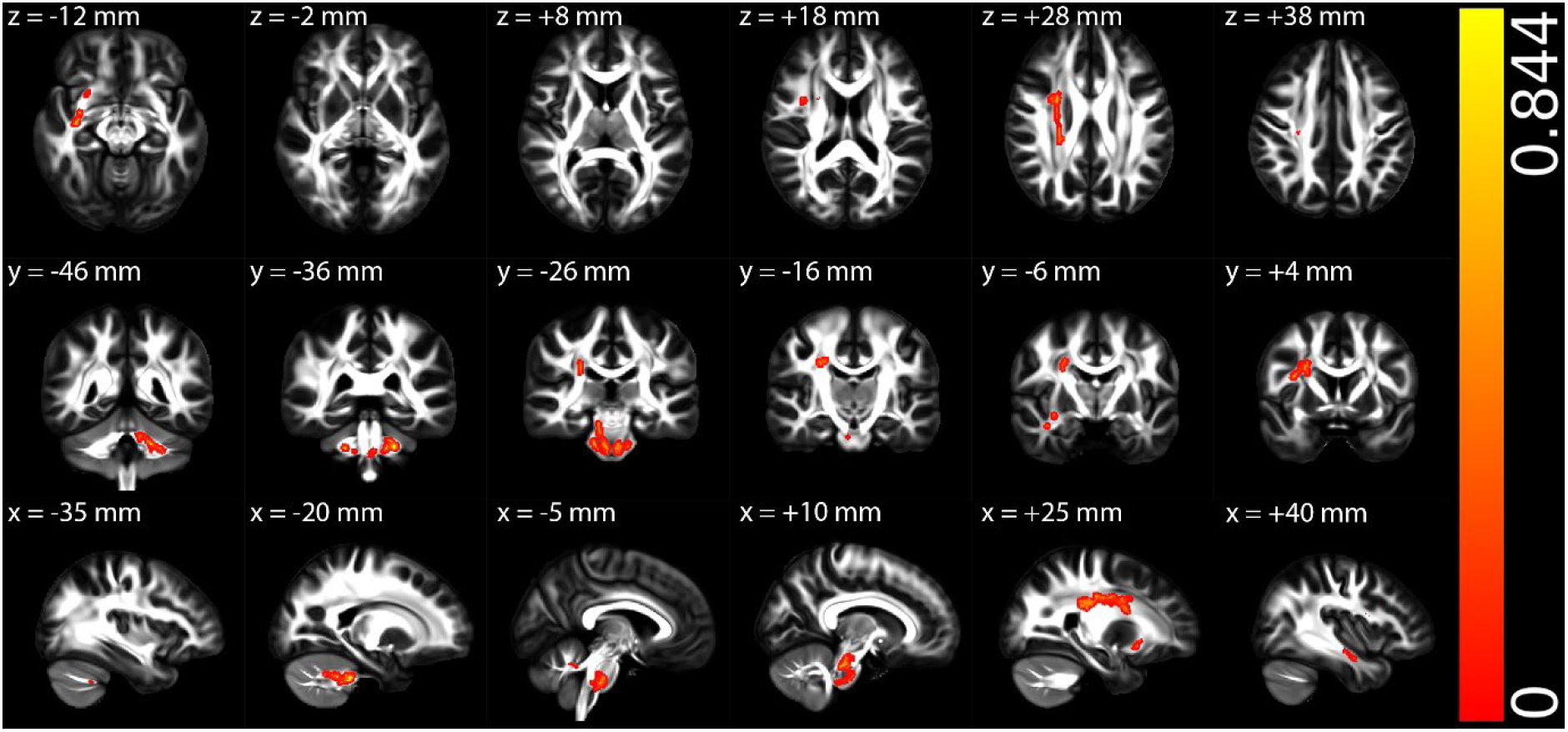
The tract-based spatial statistics (TBSS) results of axial diffusivity (AD) between the healthy control and post-infectious ME/CFS (PI-ME/CFS) patient groups in Montreal Neurological Institute (MNI) 152 standard space based on the reference FSL_HCP1065 fractional anisotropy (FA) 1×1x1mm standard-space image. The top row shows the results from six different axial slices, where the z-coordinates in MNI space from left to right are z = −12 mm, -2 mm, 8 mm, 18 mm, 28 mm, and 38 mm, respectively. The middle row shows the results from six different coronal slices, where the y-coordinates in MNI space from left to right are y = −46 mm, −36 mm, −26 mm, −16 mm, −6 mm, and 4 mm, respectively. The bottom row shows the results from six different sagittal slices, where the x-coordinates in MNI space from left to right are x = −35 mm, −20 mm, −5 mm, 10 mm, 25 mm, and 40 mm, respectively. Red-yellow clusters show the significant increased AD in PI-ME/CFS patients.

### Higher AD in PI-ME/CFS correlated with worse physical health

Among all participants in the PI-ME/CFS group, significantly negative correlations were observed between AD and physical component summary (PCS) in the following fibre tracts (Fig. 3): association fibres (right superior longitudinal fasciculus, right superior fronto-occipital fasciculus, and right external capsule), and projection fibres (right posterior thalamic radiation, right superior corona radiata, right anterior corona radiata, right posterior corona radiata, right posterior limb of internal capsule, right retrolenticular part of internal capsule, and right anterior limb of internal capsule). There were no other significant correlations between AD and three clinical scores: mental component summary (MCS), global Pittsburgh Sleep Quality Index (PSQI), and Bell’s Disability Scale (BDS) scores, for all participants in the PI-ME/CFS group.

**Fig. 3.**
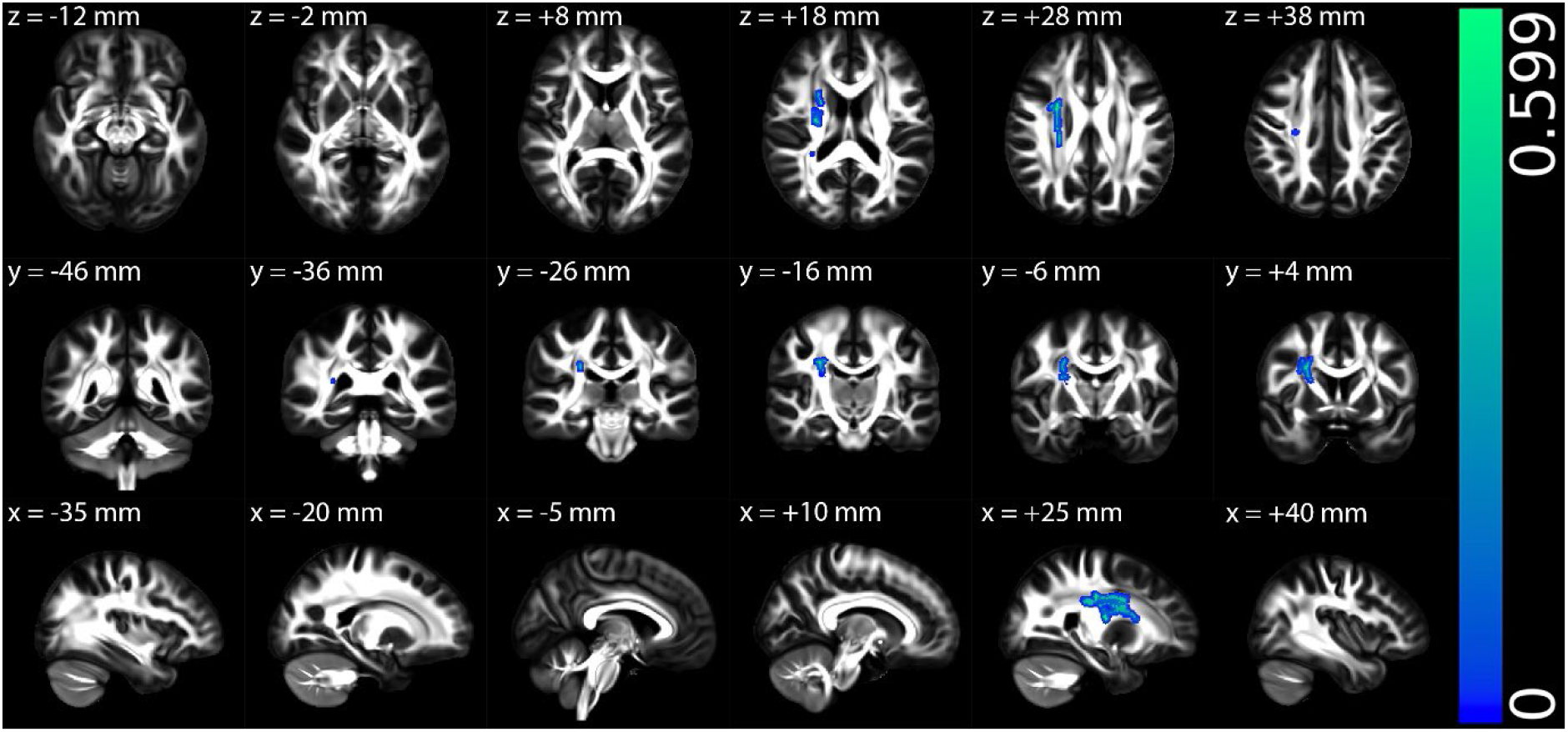
The multiple regression results of axial diffusivity (AD) with physical component summary (PCS) among all participants in the post-infectious ME/CFS (PI-ME/CFS) group in Montreal Neurological Institute (MNI) 152 standard space based on the reference FSL_HCP1065 fractional anisotropy (FA) 1×1x1mm standard-space image. The top row shows the results from six different axial slices, where the z-coordinates in MNI space from left to right are z = −12 mm, −2 mm, 8 mm, 18 mm, 28 mm, and 38 mm, respectively. The middle row shows the results from six different coronal slices, where the y-coordinates in MNI space from left to right are y = −46 mm, −36 mm, −26 mm, −16 mm, −6 mm, and 4 mm, respectively. The bottom row shows the results from six different sagittal slices, where the x-coordinates in MNI space from left to right are x = −35 mm, −20 mm, −5 mm, 10 mm, 25 mm, and 40 mm, respectively. Blue-green clusters show the significant negative correlation of AD with PCS for all participants in the PI-ME/CFS group.

For the multiple regressions exclusively conducted on the PI-ME/CFS patient group, there were no other significant correlations between AD with five clinical scores, MCS, PCS, global PSQI, BDS, and disease severity scores.

Given the lack of differences between patients and HCs in FA, MD and RD, the multiple regression analyses between those measures and the clinical measures in the PI-ME/CFS group were provided in Appendix A.1 in the supplementary information.

### Lower AD in GO-ME/CFS patients than HCs

Figure 4 illustrates the TBSS AD results between the HCs and GO-ME/CFS patient groups in MNI 152 standard space. As shown in Fig. 4, the AD measures in the GO-ME/CFS patient group were significantly lower than those in the HCs in the commissural fibres (body and genu of corpus callosum). There were no significant group differences in TBSS for FA, MD, and RD between patients and HCs in the GO-ME/CFS group.

**Fig. 4.**
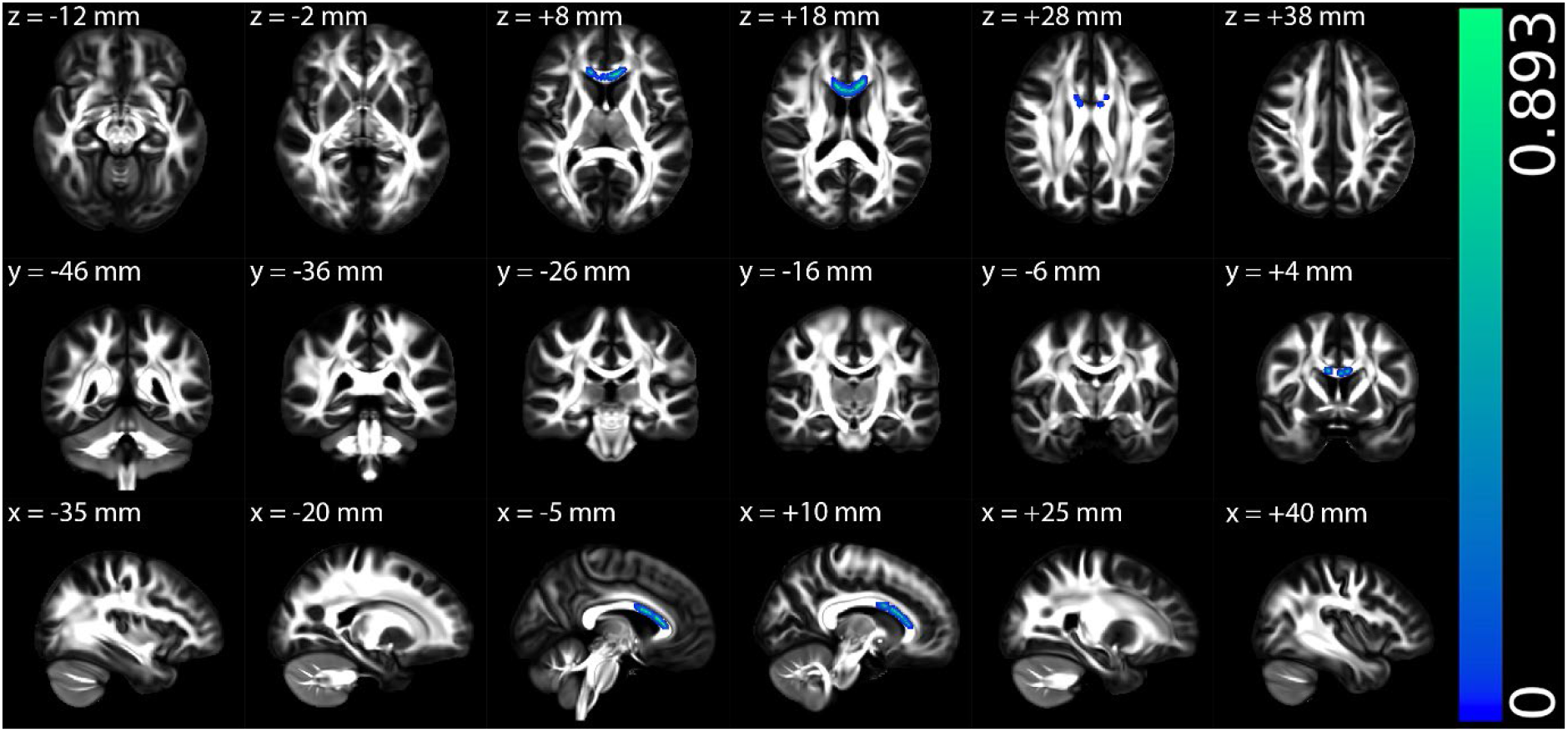
The tract-based spatial statistics (TBSS) results of axial diffusivity (AD) between the healthy control and gradual onset ME/CFS (GO-ME/CFS) patient groups in Montreal Neurological Institute (MNI) 152 standard space based on the reference FSL_HCP1065 fractional anisotropy (FA) 1×1x1mm standard-space image. The top row shows the results from six different axial slices, where the z-coordinates in MNI space from left to right are z = −12 mm, −2 mm, 8 mm, 18 mm, 28 mm, and 38 mm, respectively. The middle row shows the results from six different coronal slices, where the y-coordinates in MNI space from left to right are y = −46 mm, −36 mm, −26 mm, −16 mm, −6 mm, and 4 mm, respectively. The bottom row shows the results from six different sagittal slices, where the x-coordinates in MNI space from left to right are x = −35 mm, −20 mm, −5 mm, 10 mm, 25 mm, and 40 mm, respectively. Blue-green clusters show the significant decreased AD in GO-ME/CFS patients.

### Lower AD in GO-ME/CFS correlated with worse mental health

Among all participants in the GO-ME/CFS group, significantly positive correlations were observed between AD and MCS in the following fibre tracts (Fig. 5): commissural fibres (body and genu of corpus callosum), and projection fibres (superior corona radiata, and anterior corona radiata). There were no other significant correlations between AD with three clinical scores, PCS, global PSQI, and BDS scores, for all participants in the GO-ME/CFS group.

**Fig. 5.**
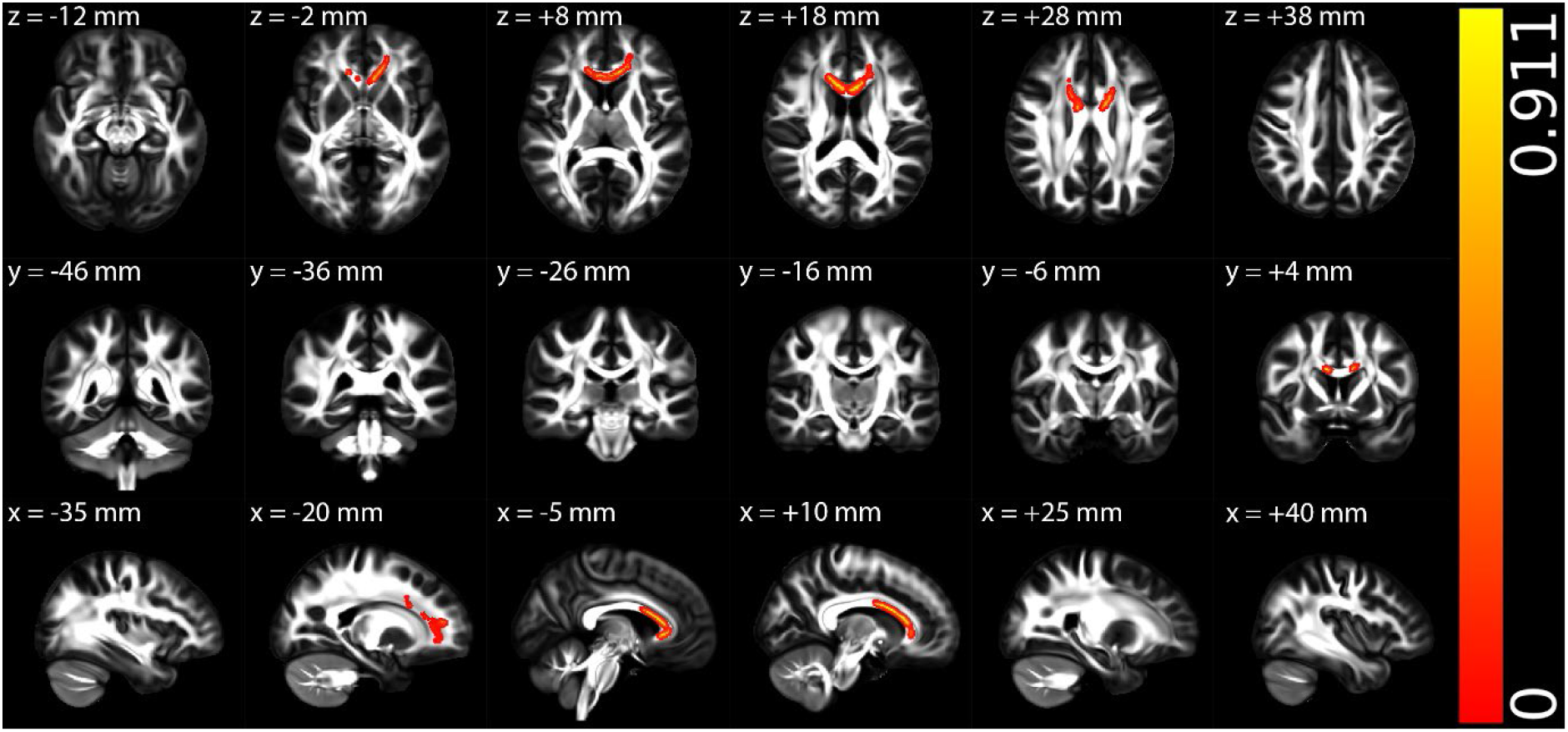
The multiple regression results of axial diffusivity (AD) with mental component summary (MCS) among all participants in the gradual onset ME/CFS (GO-ME/CFS) group in Montreal Neurological Institute (MNI) 152 standard space based on the reference FSL_HCP1065 fractional anisotropy (FA) 1×1x1mm standard-space image. The top row shows the results from six different axial slices, where the z-coordinates in MNI space from left to right are z = −12 mm, −2 mm, 8 mm, 18 mm, 28 mm, and 38 mm, respectively. The middle row shows the results from six different coronal slices, where the y-coordinates in MNI space from left to right are y = −46 mm, −36 mm, −26 mm, −16 mm, −6 mm, and 4 mm, respectively. The bottom row shows the results from six different sagittal slices, where the x-coordinates in MNI space from left to right are x = −35 mm, −20 mm, −5 mm, 10 mm, 25 mm, and 40 mm, respectively. Red-yellow clusters show the significant positive correlation of AD with MCS for all participants in the GO-ME/CFS group.

For the multiple regressions exclusively conducted on the GO-ME/CFS patient group, there were no other significant correlations between AD with five clinical scores, MCS, PCS, global PSQI, BDS, and disease severity scores.

Given the lack of differences between patients and HCs in FA, MD and RD, the multiple regression analyses between those measures and the clinical measures in the GO-ME/CFS group were provided in Appendix A.2 in the supplementary information.

## Discussion

This study investigated WM abnormalities in post-infectious (PI-ME/CFS) and gradual-onset ME/CFS (GO-ME/CFS) patients. The key findings were WM abnormalities in opposite directions: significantly increased AD among PI-ME/CFS patients compared to HCs, and significantly decreased AD among GO-ME/CFS patients compared to controls. The increased AD measures in PI-ME/CFS were correlated with worse physical health, while decreased AD values in GO-ME/CFS were related to worse mental health. To our knowledge, this is the first study to utilise DTI to explore WM microstructural alterations in ME/CFS with different disease onsets. Therefore, there is no direct comparison for consistency of results. The consistency of our findings with previous studies was discussed in the context of indirect observations.

### Increased AD in PI-ME/CFS

Our study revealed a significant increase in AD in PI-ME/CFS patients compared to the control group in multiple WM tracts, including association and projection fibres. The affected regions, such as the right superior longitudinal fasciculus, right uncinate fasciculus, and various cerebellar peduncles, are involved in cognitive functions^23,24^, emotional processing^25^, and motor control^26^. These findings align with the diverse symptomatology of ME/CFS, including cognitive impairments and motor disturbances^27,28^. Several studies have also reported exclusively increased AD in other neurological conditions associated with axonal damage, such as COVID-19^29^, mild traumatic brain injury (TBI)^30^, and retired professional football players with multiple concussions^31^. Elevated AD has been linked to axonal injury or loss of structural integrity within WM pathways^30,31^, and our findings suggest possible alterations in the integrity or arrangement of axons within specific WM tracts, as shown in Fig. 2. Increased AD has also been associated with underlying neuroinflammatory processes within the central nervous system^32,33^. Inflammation can cause tissue swelling, demyelination, and axonal damage, disrupting the organisation of axonal fibres and increasing the diffusion of water along the principal axis of the fibres^32,33^. Neuroinflammation has been observed in patients with ME/CFS^18,34–36^, COVID-19^37,38^, and TBI^39,40^. Retired professional football players with a history of multiple concussions are at an increased risk of developing long-term neurological issues, such as chronic traumatic encephalopathy^41,42^, which is associated with neuroinflammation^43^. Therefore, increased AD may serve as a potential biomarker of neuroinflammation in the PI-ME/CFS cohort, providing valuable insights into the underlying pathophysiology of these disorders.

### Decreased AD in GO-ME/CFS

In contrast, the GO-ME/CFS patients exhibited a significant decrease in AD compared to HCs in the body and genu of the corpus callosum, which are crucial for interhemispheric communication. This finding suggests that ME/CFS patients with gradual onset may have different underlying pathophysiological mechanisms compared to those with post-infectious onset. The reduced AD in these commissural fibres could potentially contribute to cognitive and information processing deficits often reported in ME/CFS patients^44,45^. Several studies have also reported exclusively decreased AD in other neurological conditions associated with axonal degeneration and demyelination, such as major depressive disorder (MDD)^46^, schizophrenia^47^, bipolar disorder (BD)^48^, obsessive-compulsive disorder (OCD)^49^, and multiple sclerosis (MS)^50^. Decreased AD has been linked to axonal degeneration or loss within WM pathways^50^, and our findings suggest potential changes in the structural or organisation of axons within WM pathways, as shown in Fig. 4. This decrease in AD indicates different pathological processes in ME/CFS patients with gradual onset, possibly related to chronic stress or prolonged disease progression impacting axonal integrity. Increased oxidative stress and impaired antioxidant defences have been reported in ME/CFS^51^, MDD^52^, schizophrenia^53^, BD^54^, OCD^55^, and MS^56^, which can result in cellular damage and impaired function in multiple organ systems, including the brain^57^. Mitochondrial abnormalities have also been observed in ME/CFS^51^, MDD^58^, schizophrenia^59^, BD^60^, OCD^61^, and MS^62^, which contribute to fatigue, cognitive impairment, and other symptoms. Additionally, alterations in neurotransmitter systems, particularly serotonin, dopamine, and glutamate, have been implicated in these conditions^63–65^. These disruptions may influence an individual’s mood, cognitive abilities, and behavioural patterns. Moreover, abnormalities in the hypothalamic-pituitary-adrenal axis, which regulates the body’s stress response, have also been observed in ME/CFS^66^, MS^67^, and various psychiatric disorders^68–70^, affecting cortisol regulation and contributing to fatigue, sleep problems, and mood fluctuations. Furthermore, sleep disturbances and altered circadian rhythms are common in these conditions^71^, affecting hormone production, cognitive function, and overall well-being. Therefore, decreased AD may serve as a potential biomarker of chronic neurodegenerative processes in the GO-ME/CFS cohort, suggesting possible axonal degeneration and demyelination.

### Opposite WM abnormalities correlated with different aspects of healthy scores

When exploring the correlation between AD and clinical symptoms for the PI-ME/CFS group, we found a significant negative association between AD and PCS score in various WM tracts, including association and projection fibres, across all participants. This negative relationship implies that higher AD is associated with poorer physical health outcomes. This finding could reflect the impact of PI-ME/CFS on physical functioning, where increased AD may serve as a potential biomarker for disease severity in physical domains. Motor disturbances are a significant and often debilitating aspect of ME/CFS^22,78–80^. Recent studies on COVID-19 have also found motor disturbances and WM abnormalities in patients^81–83^. The observed WM changes in the PI-ME/CFS group appear to be more closely associated with the physical health aspects of the disease, rather than the mental health component, sleep quality, disability level, or disease severity. This indicates that these WM alterations may be particularly relevant to the physical functioning symptoms experienced by patients, reflecting the impact of neuroinflammation on physical capabilities in post-infectious ME/CFS patients.

For the GO-ME/CFS group, we observed a significant positive correlation between AD and MCS score in commissural fibres (body and genu of the corpus callosum) and projection fibres (superior corona radiata, anterior corona radiata) among all participants. This suggests that in ME/CFS patients with gradual onset, lower AD values (potentially indicating axonal degeneration and demyelination) in these regions are associated with worse mental health outcomes. The commissural fibres of the corpus callosum primarily enable communication and integration between the brain’s hemispheres, while the projection fibres of the corona radiata are essential for transmitting information between cortical and subcortical regions, significantly contributing to motor control, sensory processing, and various cognitive functions^84,85^. The multiple regression results suggest a potential link between WM axonal damage and the severity of cognitive and psychological symptoms in GO-ME/CFS patients. Cognitive dysfunction is a prevalent characteristic of ME/CFS, including deficits in areas such as attention, memory, and information processing speed^44,45^. Recent studies on MDD^86^, schizophrenia^87^, BD^48^, OCD^88^, and multiple sclerosis^89^ also found cognitive impairment and WM abnormalities in patients. The observed WM changes in the GO-ME/CFS group appear to be more closely associated with the mental health aspects of the disease. This indicates that these WM alterations may be particularly relevant to the cognitive and affective symptoms experienced by ME/CFS patients with gradual onset. Conducting longitudinal studies and integrating findings with other neuroimaging data, clinical information, and neuropsychological assessments could lead to a more comprehensive understanding of the biological mechanisms driving these observations.

### Biological implications

The divergent findings between the PI-ME/CFS and GO-ME/CFS groups highlight the heterogeneity of ME/CFS and underscore the importance of considering disease onset in neuroimaging studies. Increased AD in PI-ME/CFS patients might be related to acute inflammatory responses following an infection, whereas decreased AD in GO-ME/CFS patients may reflect chronic neurodegenerative processes. The clinical correlations observed, particularly the negative relationship between AD and PCS in PI-ME/CFS and the positive relationship between AD and MCS in GO-ME/CFS, suggest potential pathways for targeted therapeutic interventions. These findings also suggest a personalised approach to treating ME/CFS, considering the onset and progression of the disease. By focusing on strategies that alleviate axonal damage, reduce neuroinflammation, or enhance WM repair, we may mitigate symptoms experienced by ME/CFS patients and improve their well-being.

### Inconsistent observations of DTI indices in ME/CFS

Interestingly, we did not find significant differences in FA, MD, or RD between PI-ME/CFS patients, GO-ME/CFS patients and controls, which contrasts with previous studies reporting changes in these diffusion metrics in ME/CFS^15–17^. We hypothesise that inconsistencies in findings across studies may arise from differences in sample characteristics, patient criteria, or statistical methodologies. Table 3 highlights the key differences between our study and three previous studies^15–17^ that reported alterations in DTI metrics in ME/CFS.

**Table 3:**
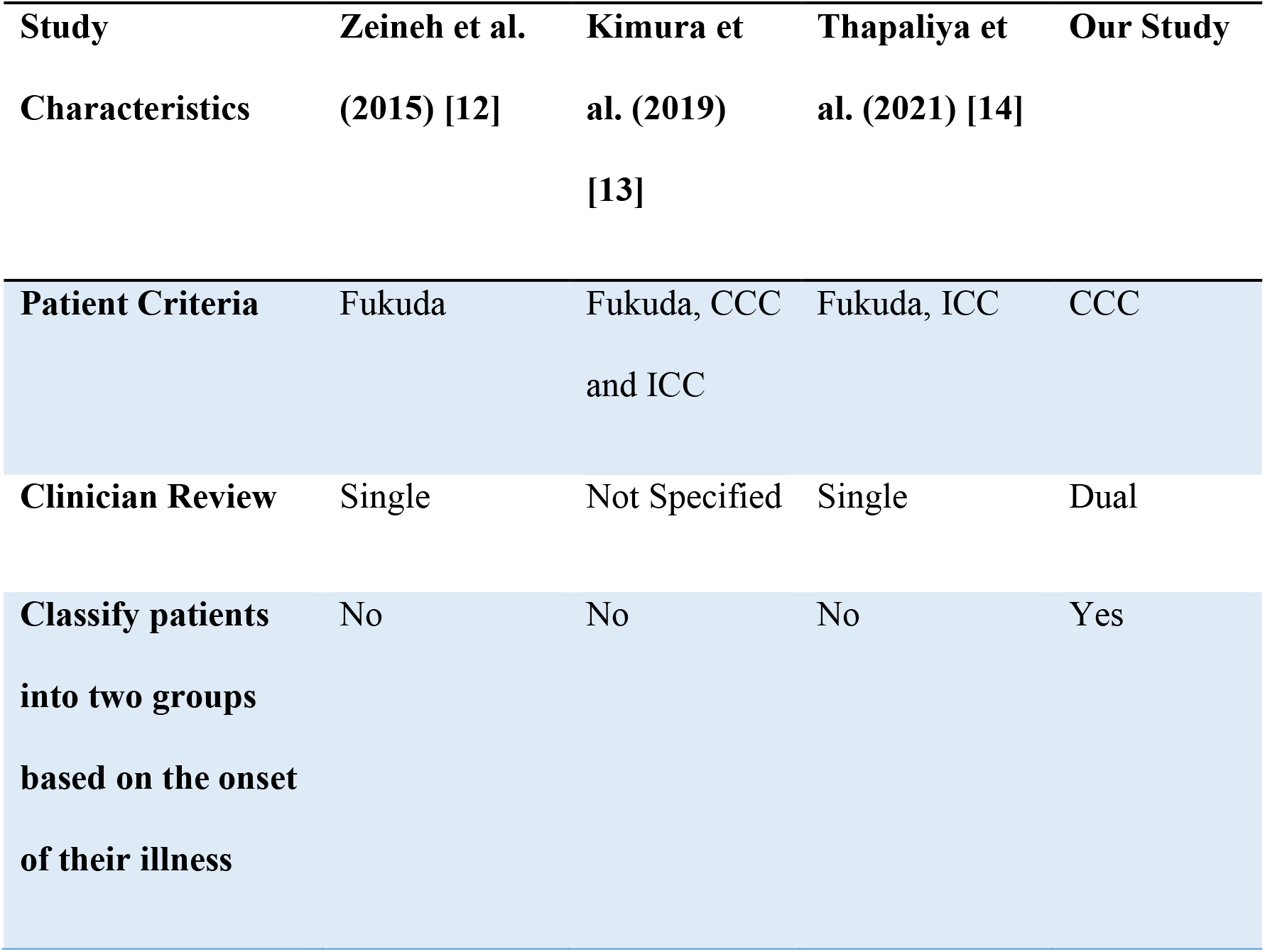
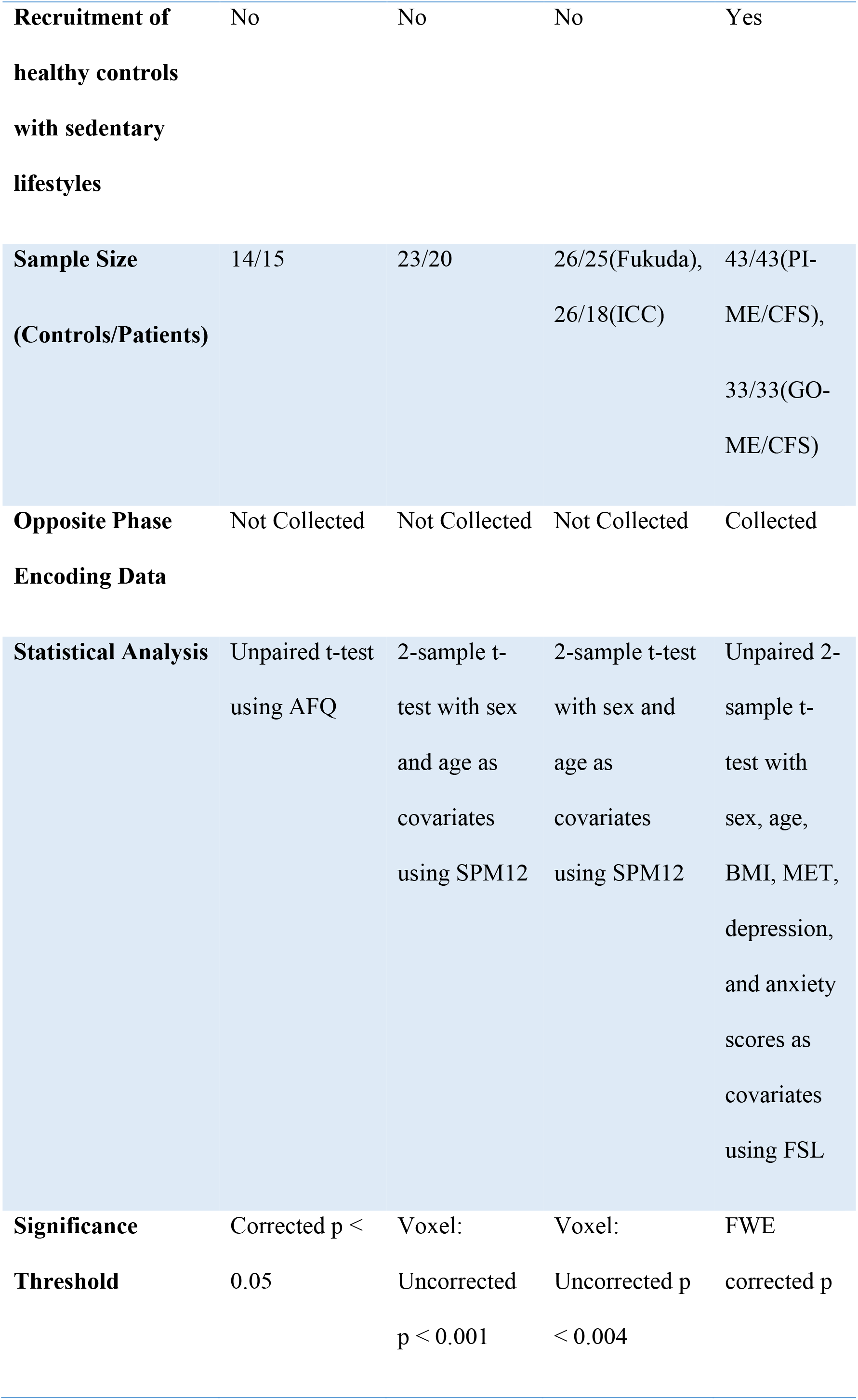
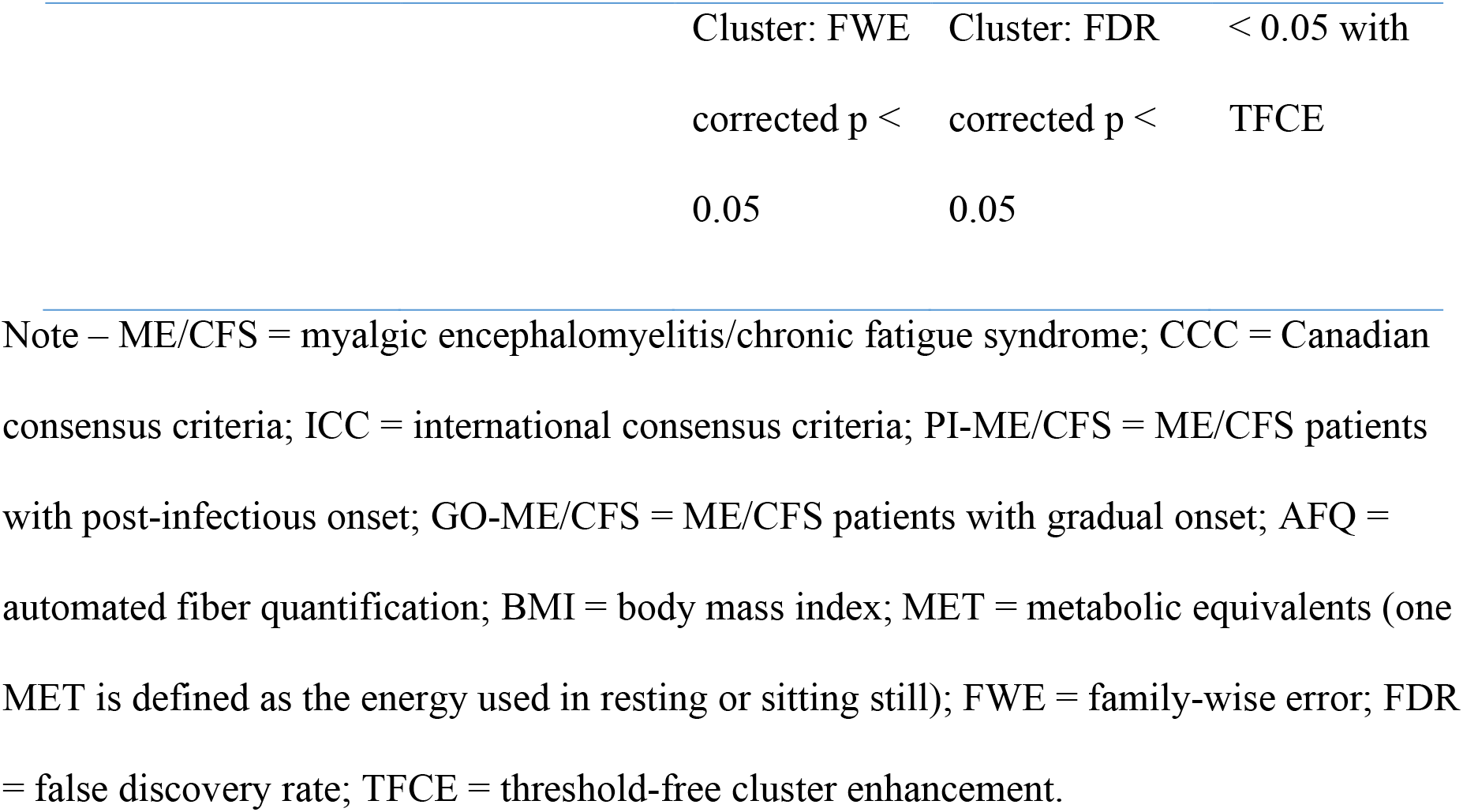
The key differences between our study and three previous studies that reported changes in diffusion tensor metrics in ME/CFS.

ME/CFS patients in our study were reviewed by two clinicians (RK and PDF) using the Canadian Consensus Criteria (CCC) Criteria^90^ to minimise the risk of including an ill-defined patient cohort. Patients were classified into two subgroups (PI-ME/CFS and GO-ME/CFS) based on the onset of their illness^1,6^ to ensure more homogeneous patient groups. Additionally, our study recruited HCs with sedentary lifestyles to mitigate the deconditioning effects of ME/CFS patients who have reduced physical activities^22^. Furthermore, our study has an appropriate sample size with equal numbers of patients and controls in the PI-ME/CFS group (N = 86) and the GO-ME/CFS group (N = 66). While unequal group size can be addressed by statistical adjustments, equal group sizes increase the power of the analysis, enhancing the likelihood of detecting true differences between groups^72^. Equal group sizes also reduce biases related to differences in experimental units among groups, leading to more accurate and reliable results^73^.

Our study included additional opposite phase encoding data to enhance the correction of eddy current distortions, which may contribute to more accurate DTI results^74^. Furthermore, our statistical analysis employed a comprehensive statistical approach with a conservative significance threshold [family-wise error (FWE) corrected p-value < 0.05 with threshold-free cluster enhancement (TFCE)]. While FWE correction reduces the likelihood of false positive results, it may be less sensitive in detecting true effects than false discovery rate (FDR) correction^75,76^. The TFCE method, however, can offer better sensitivity than cluster-based thresholding^77^.

### Limitations

This study has some limitations that should be acknowledged. Firstly, we focused on WM fibre characteristics as part of a multimodal MRI investigation of ME/CFS. We observed increased ADs in the PI-ME/CFS group associated with poorer physical summary scores, and decreased ADs in the GO-ME/CFS group related to worse mental summary scores. However, WM fibre measures based on AD were not associated with sleep qualities, disability level, or disease severity. These results suggested that ME/CFS may be heterogeneous, and other brain aspects might also be impaired. Future research will explore additional brain characteristics and their relationships with symptoms. Secondly, the significant correlations between AD and PCS in the PI-ME/CFS group and AD and MCS in the GO-ME/CFS group were lost when we conducted the multiple regression analyses exclusively on the patient group. Our study did not account for potential confounding factors, such as medication usage, comorbidities, or disease duration. These factors could have affected the observed relationships between AD and clinical measures, particularly within the patient group. The increased heterogeneity in AD and clinical measures among patients, along with the smaller sample size of the patient group compared to the combined group, may have contributed to the loss of significant correlations when analysing patients separately. Future studies should aim to control for these variables or investigate their potential moderating effects on the relationship between WM integrity and ME/CFS.

In summary, this study provides significant insights into the WM alterations in ME/CFS patients, distinguishing between those with post-infectious (PI-ME/CFS) and gradual onset (GO-ME/CFS) subtypes. The findings reveal distinct patterns of changes in AD between these subgroups: PI-ME/CFS patients exhibit increased AD in various association and projection fibres, potentially reflecting post-infectious inflammatory processes, whereas GO-ME/CFS patients show decreased AD in commissural fibres, suggesting possible axonal degeneration. These results indicate different underlying pathophysiological mechanisms between the two ME/CFS subtypes and suggest that AD may serve as a sensitive indicator of WM pathology.

## Methods

The study was approved by the University of the Sunshine Coast Ethics Committee (A191288). Participants provided written consent prior to taking part in the study. All datasets were de-identified before processing.

The inclusion and exclusion criteria for the participants are detailed in our protocol paper^21^. Adult participants aged 18 to 65 years were recruited. Critical exclusion criteria include individuals (1) out of the age range, (2) with a BMI >35, and (3) who smoke (including marijuana or substance usage). Additional exclusion criteria for HCs included individuals with mental disorders. ME/CFS patients were reviewed by two clinicians (RK and PDF) using the CCC Criteria and classified into two groups based on the onset of their illness. The PI-ME/CFS group was defined as those whose ME/CFS symptoms manifested immediately following a virus or bacterial infection, and the GO-ME/CFS group was defined as those whose ME/CFS symptoms developed gradually over several months without clearly identified triggers. This study also specifically recruited healthy control subjects with sedentary lifestyles (< 60 minutes of moderate or high-intensity activity (i.e., exercise) per week)^91^ to minimise the confounding effects of disease-related deconditioning. Activity level for all participants were monitored using the Actigraph GT3X-BT device (ActiGraph LLC., United States) for 14 days, including seven days before and after MRI scans.

### Clinical measures

The Hospital Anxiety and Depression Scale (HADS)^92^, 36-item Short-Form (SF-36) Health Survey^93^, Pittsburgh Sleep Quality Index (PSQI)^94^, and Bell’s Disability Scale (BDS)^95^ questionnaires were completed by all participants. The HADS-depression score, HADS-anxiety score, SF-36 mental component summary (MCS), SF-36 physical component summary (PCS), global PSQI score, and BDS score were extracted to assess participants’ symptoms. Additionally, the disease severity of ME/CFS patients was assessed by two clinicians (RK and PDF). Furthermore, the MET of the task from the Actigraph data was calculated using the Freedson Adult (1998) algorithm^96^ via ActiLife® software to measure activity for each participant.

Some behavioural measures of MET, depression, and anxiety scores were missing during data collection. To address these gaps, we adopted an imputation approach based on age-specific groups^97^. For seven MET measures in HCs, we imputed missing values using the mean of their corresponding age-specific control group, which had a range of 10 years between ages 18 and 65. Similarly, for one PI-ME/CFS and three GO-ME/CFS patients, missing MET values were imputed using the mean of the corresponding 10-year age-specific ME/CFS patient group. Additionally, missing values for depression and anxiety scores for one control participant were imputed using the mean of the corresponding 10-year age-specific control group. This systematic use of 10-year age-specific group means to impute missing values helped maintain the integrity of the dataset and preserved the relationships between these variables and others included in the analysis.

### MRI acquisition

Brain images were acquired using a 3T Skyra MRI scanner (Germany, Erlangen) with a 64- channel head coil. The structural MRI data were collected using a T1-weighted magnetisation prepared rapid gradient-echo sequence (MPRAGE): 208 slices, dimension 256 × 256, voxel size 1mm × 1mm × 1mm, TR/TE = 2,200/1.71ms, flip angle 7°. The DTI data were acquired using a multiband EPI sequence (72 slices, multiband factor 3, dimension = 114 × 114, voxel size 2 × 2 × 2 mm^3^, TR/TE = 4,500/123ms; free diffusion mode, bipolar diffusion scheme with two diffusion weighting of b = 1000 and b = 2,500 s/mm^2^ with noncolinear diffusion directions 27 and 62, respectively, and eight volumes of b = 0 s/mm^2^, and phase encoding direction = anterior to posterior,). Six volumes of b = 0 s/mm^2^ with opposite phase encoding direction in posterior to anterior are also collected for the eddy current correction.

### Imaging processing

The DTI data were analysed using the established tool, TBSS^98^ in the Functional MRI of the Brain (FMRIB) Software Library v6.0 (FSL)^99^. TBSS is a whole-brain approach that allows for the analysis of diffusion characteristics across the entire brain.

The standard FSL FDT (FMRIB’s Diffusion Toolbox)^100^ pipeline was used to preprocess the diffusion data. The effects of motion for the six volumes of b = 0 s/mm^2^ with opposite phase-encoding direction EPI images were reduced using ‘mcflirt’^101^. The warping field required for EPI distortion correction was calculated using ‘TOPUP’^74^. The brain mask was generated using ‘bet2’^102^ with threshold 0.2 on the b = 0 s/mm^2^ images, which were outputs from the ‘TOPUP’ command. Eddy current induced distortion correction and motion correction were performed using ‘eddy_openmp’^103,104^, and the eddy quality control (qc) tool^105^ was used to perform quality control at both single subject and group level. Diffusion metrics of FA, MD, AD and RD, were created by fitting a diffusion tensor model at each voxel using ‘dtifit’. Each participant’s FA image was nonlinearly registered into MNI 152 standard space using TBSS. The FSL_HCP1065 FA 1×1x1mm standard-space image was used as the target. The same nonlinear deformations were also applied to other non-FA images (MD, AD and RD).

### Statistical analysis

#### Demographic data and behavioural scores

The normal distribution of the demographic data and behavioural scores were tested using the Lilliesfors test^106^. If any data do not exhibit a normal distribution, the Wilcoxon rank-sum test^107^ (non-parametric test) was used to compare the data between groups.

#### TBSS analysis

The unpaired two-sample t-test was implemented using the ‘randomise’ tool^108^ in FSL to compare the differences in each diffusion metric between the healthy control and patient groups. Sex, age, BMI, MET, depression, and anxiety scores were included as nuisance covariates in TBSS analyses. This comparison was conducted using a nonparametric permutation inference method with 10,000 permutations at each voxel^109^. FWE corrected p < 0.05 with TFCE was used to indicate significant differences.

#### Multiple regression

To further explore the relationship between microstructural changes and clinical symptoms, multiple regression analyses were performed using the ‘randomise’ tool. Multiple regressions between DTI metrics with four clinical scores, MCS, PCS, global PSQI, and BDS scores, were performed for all participants. The same multiple regression analyses were also conducted exclusively on the patient group. Additionally, we examined the associations between DTI metrics and disease severity measures, but this analysis was limited to the patient group only. Note that one GO-ME/CFS participant was excluded from multiple regression analyses for MCS and PCS due to missing scores, while one GO-ME/CFS and two PI-ME/CFS participants were excluded from analyses for global PSQI due to missing scores. All multiple regressions included sex, age, BMI, MET, depression, and anxiety scores as nuisance covariates. FWE corrected p < 0.05 with TFCE was used to reflect significant regression results.

## Data availability

The demographics, symptom scores, and preprocessed DTI metrics (FA, MD, AD and RD) of each participant are available in the Zenodo repository (DOI:10.5281/zenodo.12791948). Source data are provided with this paper.

## Code availability

The standard analysis pipeline of FSL was used, https://fsl.fmrib.ox.ac.uk/fsl/fslwiki/FDT/UserGuide.

## Data Availability

The demographics, symptom scores, and preprocessed DTI metrics (FA, MD, AD and RD) of each participant are available in the Zenodo repository (DOI:10.5281/zenodo.12791948).

https://doi.org/10.5281/zenodo.12791948

## Acknowledgements

The study was supported by the National Health and Medical Research Council of Australia (NHMRC) Ideas Grant Scheme (GNT1184219) and The Mason Foundation (MAS2018F00024). We are also thankful to all participants who were involved in this study.

## Author Contributions

Q.Y., R.A.K., P.D., V.D.C., and Z.Y.S. conceived the project; Q.Y. and Z.Y.S. designed the study; Q.Y., A.B., and Z.Y.S. performed data analysis; Q.Y., R.A.K., P.D., V.D.C., G.A.B., T.Y., and Z.Y.S. interpreted the results; Q.Y. drafted the manuscript, and all authors contributed to editing it.

## Supplementary Information

### Appendix A

The appendix A shows the results for the multiple regression analyses between fractional anisotropy (FA), mean diffusivity (MD), radial diffusivity (RD), and the clinical measures for participants included in the final analysis for post-infectious myalgic encephalomyelitis/chronic fatigue syndrome (PI-ME/CFS) (43 PI-ME/CFS and 43 healthy controls (HCs)) and gradual onset ME/CFS (GO-ME/CFS) (33 GO-ME/CFS and 33 HCs) studies.

#### A. Multiple regression with clinical measures in the PI-ME/CFS group

Among all participants in the PI-ME/CFS group, there were no other significant correlations between FA, MD, and RD with four clinical scores, mental component summary (MCS), physical component summary (PCS), global Pittsburgh Sleep Quality Index (PSQI), and Bell’s Disability Scale (BDS) scores.

For the multiple regressions exclusively conducted on the PI-ME/CFS patient group, significantly negative correlations were observed between FA and MCS in the following fibre tracts (Supplementary Fig. 1): the cerebral white matter (WM) tracts (superior longitudinal fasciculus, cingulum cingulate, and left posterior thalamic radiation), the cerebral peduncles and radiations (superior corona radiata, anterior corona radiata, and posterior corona radiata), and the cerebral commissures (corpus callosum and left tapetum). In addition, significantly positive correlations were also observed between RD and MCS in the following fibre tracts (Supplementary Fig. 2): the cerebral WM tracts (superior longitudinal fasciculus, left cingulum cingulate, left superior fronto-occipital fasciculus, and left posterior thalamic radiation), the cerebral peduncles and radiations (superior corona radiata, anterior corona radiata, and left posterior corona radiata), the internal capsule (left posterior limb of internal capsule and left anterior limb of internal capsule), the external capsule, and the cerebral commissures (corpus callosum and left tapetum).

There were no other significant correlations between FA, MD, and RD with five clinical scores, MCS, PCS, global PSQI, BDS, and disease severity scores, for the multiple regressions exclusively conducted on the PI-ME/CFS patient group.

#### B. Multiple regression with clinical measures in the GO-ME/CFS group

Among all participants in the GO-ME/CFS group, significantly positive correlations were observed between FA and MCS in the following fibre tracts (Supplementary Fig. 3): the cerebral peduncles and radiations (left superior corona radiata and anterior corona radiata), and the cerebral commissures (body and genu of corpus callosum).

There were no other significant correlations between FA, MD, and RD with four clinical scores, MCS, PCS, global PSQI, and BDS scores, for all participants in the GO-ME/CFS group.

For the multiple regressions exclusively conducted on the GO-ME/CFS patient group, significantly positive correlations were observed between RD and disease severity in the following fibre tracts (Supplementary Fig. 4): the cerebral WM tracts (superior longitudinal fasciculus, cingulum cingulate, uncinate fasciculus, superior fronto-occipital fasciculus, and posterior thalamic radiation), the cerebral peduncles and radiations (cerebellar peduncle, superior corona radiata, anterior corona radiata, and posterior corona radiata), the internal capsule (posterior limb of internal capsule, retrolenticular part of internal capsule, anterior limb of internal capsule, and sagittal stratum), the external capsule, the cerebral commissures (corpus callosum and tapetum), and limbic system (stria terminalis). Furthermore, Supplementary Fig. 5 shows the observed significantly negative correlations between FA and disease severity for GO-ME/CFS patient group. Except for the regions as shown in Supplementary Fig. 4 for the significantly positive correlations were observed between RD and disease severity, the FA revealed widespread significant negative correlation with disease severity in the following regions: the cerebral WM tracts (cingulum hippocampus), the cerebral peduncles and radiations (inferior cerebellar peduncle, middle cerebellar peduncle, and superior cerebellar peduncle), brain stem (corticospinal tract, and medial lemniscus), and limbic system (fornix).

There were no other significant correlations between FA, MD, and RD with five clinical scores, MCS, PCS, global PSQI, BDS, and disease severity scores, for the multiple regressions exclusively conducted on the GO-ME/CFS patient group.

### Appendix B

The appendix B shows the results for all eligible participants for PI-ME/CFS (43 PI-ME/CFS and 58 HCs) and GO-ME/CFS (33 GO-ME/CFS and 66 HCs) groups before age, sex, metabolic equivalent rate (MET) and number matching (1:1).

#### B.1. Participant characteristics

Supplementary Table 1 presents the demographic characteristics and behavioural assessments for all eligible participants for PI-ME/CFS before age, sex, MET and number matching (1:1). There is no significant difference in age (P = 0.07), body mass index (BMI) (P = 0.48). and MET rate (P = 0.07) between the PI-ME/CFS patients and healthy control groups. Compared to PI-ME/CFS, the healthy control group showed reduced Hospital Anxiety and Depression Scale (HADS) anxiety (P < 0.001) and depression (P < 0.001) and increased 36-item Short- Form (SF-36) mental health (P < 0.001) and physical health (P < 0.001). The healthy control group also have better overall sleep quality (P < 0.001) and lower level of disability (P < 0.001) than the patient’s group.

Supplementary Table 2 demonstrates the demographic characteristics and behavioural assessments for all eligible participants for GO-ME/CFS before age, sex, MET and number matching (1:1). There is no significant difference in age (P = 0.10), BMI (P = 0.86). and MET rate (P = 0.07) between the GO-ME/CFS patients and healthy control groups. Compared to GO-ME/CFS, the healthy control group showed reduced HADS anxiety (P < 0.001) and depression (P < 0.001) and increased SF-36 mental health (P < 0.001) and physical health (P < 0.001). The healthy control group also have better overall sleep quality (P < 0.001) and lower level of disability (P < 0.001) than the patient’s group.

#### B.2. TBSS group comparison in the PI-ME/CFS group

Supplementary Fig. 6 illustrates the tract-based spatial statistics (TBSS) axial diffusivity (AD) results between the healthy control and PI-ME/CFS patient groups in Montreal Neurological Institute (MNI) 152 standard space. As shown in Supplementary Fig. 6, the AD in the PI- ME/CFS patient group were significantly higher than those in the healthy control in the following fibre tracts: the cerebral WM tracts (right superior longitudinal fasciculus, uncinate fasciculus, cingulum hippocampus, right superior fronto-occipital fasciculus, and posterior thalamic radiation), the cerebral peduncles and radiations (cerebellar peduncle, inferior cerebellar peduncle, middle cerebellar peduncle, superior cerebellar peduncle, right superior corona radiata, anterior corona radiata, and posterior corona radiata), the internal capsule (right posterior limb of internal capsule, retrolenticular part of internal capsule, right anterior limb of internal capsule, and sagittal stratum), the external capsule, the cerebral commissures (body and splenium of corpus callosum, and right tapetum), brain stem (corticospinal tract and medial lemniscus), and limbic system (stria terminalis).

There were no significant group differences in TBSS for FA, MD, and RD for all eligible participants in PI-ME/CFS group.

#### B.3. Multiple regression with clinical measures in the PI-ME/CFS group

Among all eligible participants in the PI-ME/CFS group, significantly negative correlations were observed between AD and PCS in the following fibre tracts (Supplementary Fig. 7): the cerebral WM tracts (right superior longitudinal fasciculus, right superior fronto-occipital fasciculus, and right posterior thalamic radiation), the cerebral peduncles and radiations (right superior corona radiata, right anterior corona radiata, and right posterior corona radiata), the internal capsule (right posterior limb of internal capsule, right retrolenticular part of internal capsule, right anterior limb of internal capsule, and right sagittal stratum), the right external capsule, and the cerebral commissures (right tapetum). In addition, Supplementary Fig. 8 demonstrates the observed significantly negative correlations between AD and BDS among all eligible participants in the PI-ME/CFS group. Except for the regions as shown in Supplementary Fig. 7 for the significantly negative correlations were observed between AD and PCS, the AD revealed widespread significant negative correlation with BDS in the right stria terminalis.

There were no other significant correlations between DTI metrics (FA, MD, AD and RD) with four clinical scores, MCS, PCS, PSQI, and BDS scores, for all eligible participants in the PI- ME/CFS group.

#### B.4. TBSS group comparison in the GO-ME/CFS group

Supplementary Fig. 9 illustrates the TBSS AD results between the healthy control and GO- ME/CFS patient groups in MNI 152 standard space. As shown in Supplementary Fig. 9, the AD in the GO-ME/CFS patient group were significantly lower than those in the healthy control in the cerebral commissures (body and genu of corpus callosum). In addition, Supplementary Figs. S10 and S11 show the observed significantly decreased MD and RD in the GO-ME/CFS patient group. Except for the regions as shown in Supplementary Fig. 9 for the significantly decreased AD in the patients, the MD and RD revealed widespread significant decreased regions in the cerebral peduncles and radiations (superior corona radiata, anterior corona radiata, and left posterior corona radiata).

There was no significant group difference in TBSS for FA for all eligible participants in the GO-ME/CFS group.

#### B.5. Multiple regression with clinical measures in the GO-ME/CFS group

Among all eligible participants in the GO-ME/CFS group, there were no significant correlations between any of the DTI metrics (FA, MD, AD and RD) and four clinical measures (MCS, PCS, global PSQI, and BDS scores.

---

**Supplementary Fig. 1.**
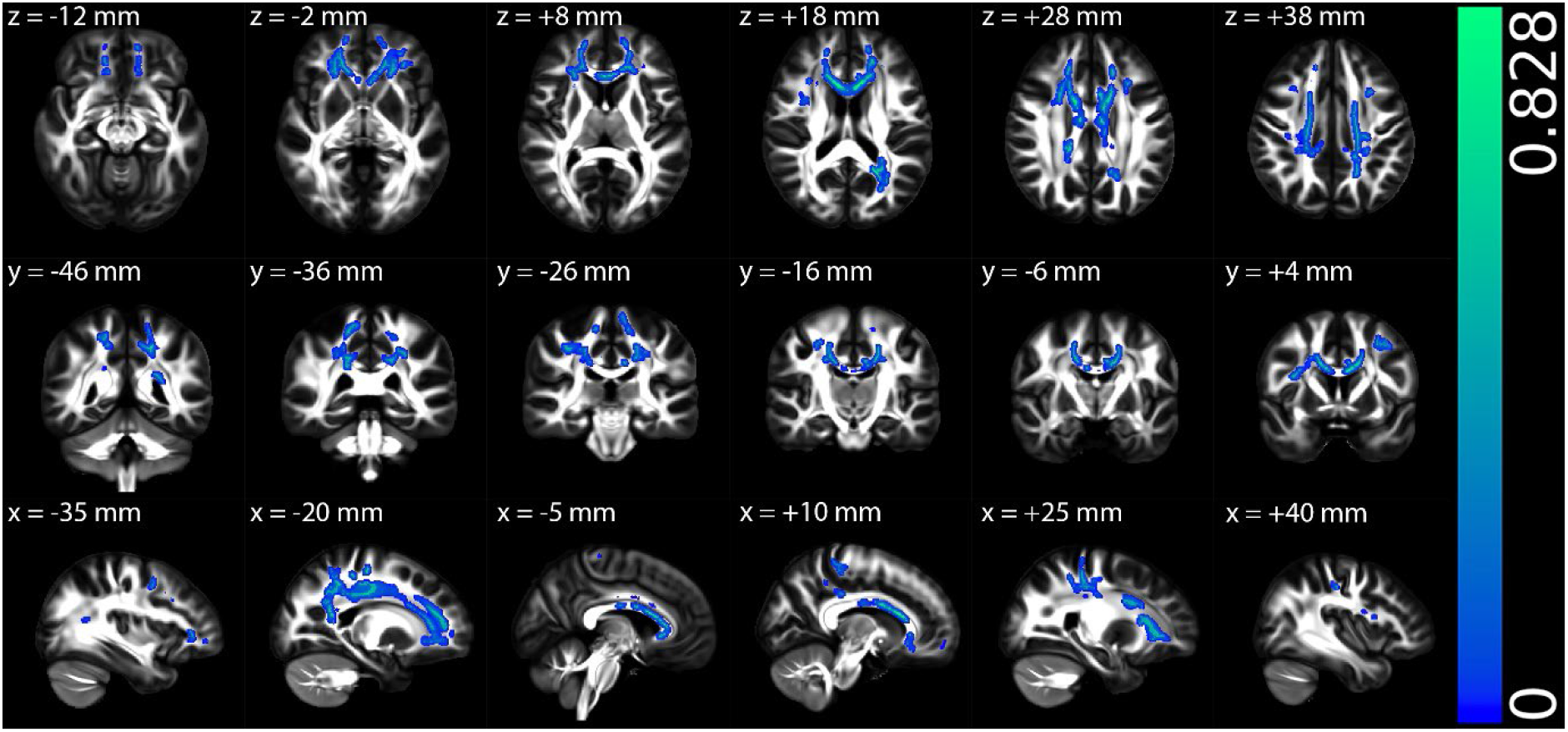
The multiple regression results of fractional anisotropy (FA) with mental component summary (MCS) for post-infectious ME/CFS (PI-ME/CFS) patient group in Montreal Neurological Institute (MNI) 152 standard space based on the reference FSL_HCP1065 FA 1×1x1mm standard-space image. The top row shows the results from six different axial slices, where the z-coordinates in MNI space from left to right are z = −12 mm, −2 mm, 8 mm, 18 mm, 28 mm, and 38 mm, respectively. The middle row shows the results from six different coronal slices, where the y-coordinates in MNI space from left to right are y = −46 mm, −36 mm, −26 mm, −16 mm, −6 mm, and 4 mm, respectively. The bottom row shows the results from six different sagittal slices, where the x-coordinates in MNI space from left to right are x = −35 mm, −20 mm, −5 mm, 10 mm, 25 mm, and 40 mm, respectively. Blue-green clusters show the significant negative correlation of FA with MCS in the PI-ME/CFS patient group.

**Supplementary Fig. 2.**
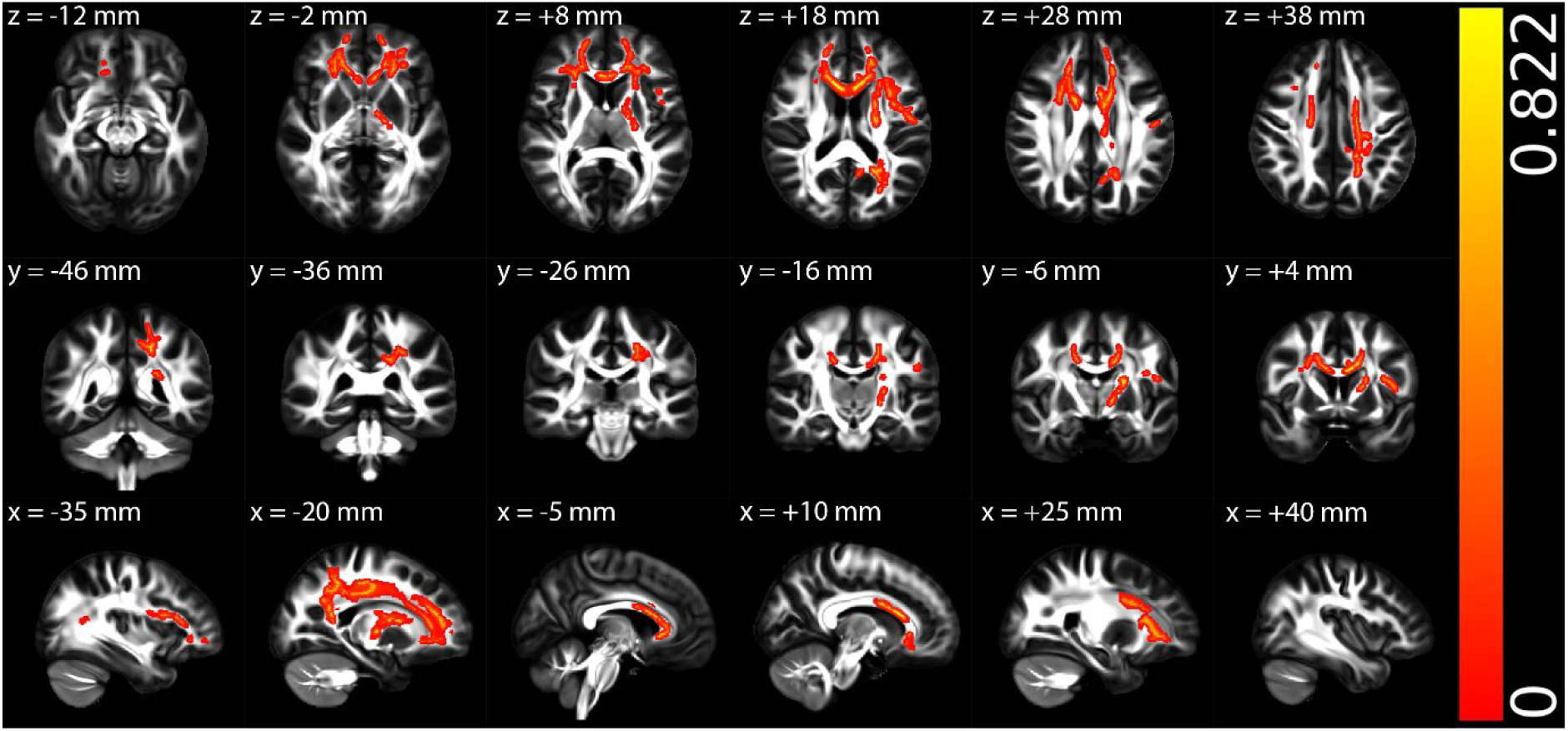
The multiple regression results of radial diffusivity (RD) with mental component summary (MCS) in the post-infectious ME/CFS (PI-ME/CFS) patient group in Montreal Neurological Institute (MNI) 152 standard space based on the reference FSL_HCP1065 FA 1×1x1mm standard-space image. The top row shows the results from six different axial slices, where the z-coordinates in MNI space from left to right are z = −12 mm, −2 mm, 8 mm, 18 mm, 28 mm, and 38 mm, respectively. The middle row shows the results from six different coronal slices, where the y-coordinates in MNI space from left to right are y = −46 mm, −36 mm, −26 mm, −16 mm, −6 mm, and 4 mm, respectively. The bottom row shows the results from six different sagittal slices, where the x-coordinates in MNI space from left to right are x = −35 mm, −20 mm, −5 mm, 10 mm, 25 mm, and 40 mm, respectively. Red-yellow clusters show the significant positive correlation of RD with MCS in the PI-ME/CFS patient group.

**Supplementary Fig. 3.**
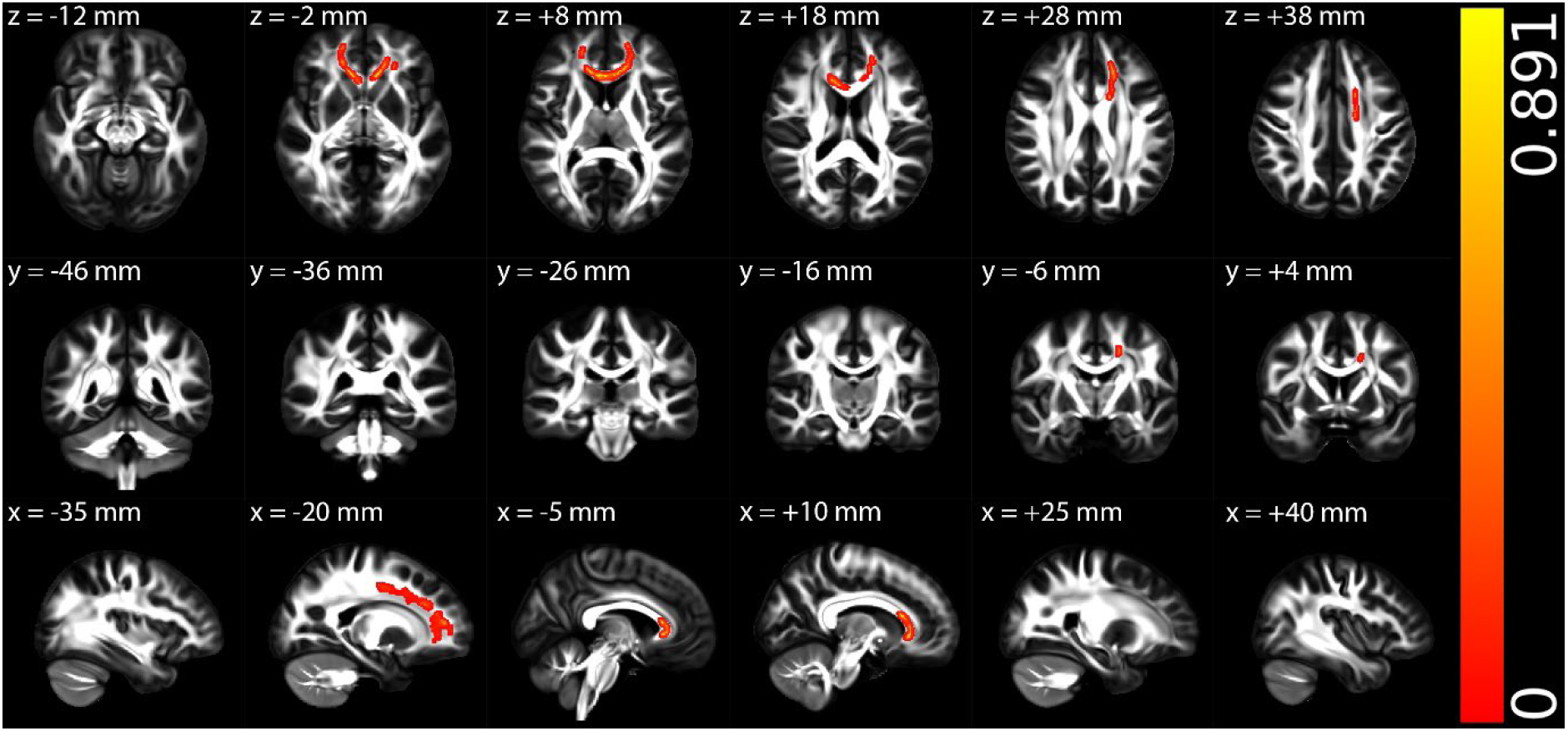
The multiple regression results of fractional anisotropy (FA) with mental component summary (MCS) among all participants in the gradual onset ME/CFS (GO- ME/CFS) group in Montreal Neurological Institute (MNI) 152 standard space based on the reference FSL_HCP1065 fractional anisotropy (FA) 1×1x1mm standard-space image. The top row shows the results from six different axial slices, where the z-coordinates in MNI space from left to right are z = −12 mm, −2 mm, 8 mm, 18 mm, 28 mm, and 38 mm, respectively. The middle row shows the results from six different coronal slices, where the y-coordinates in MNI space from left to right are y = −46 mm, −36 mm, −26 mm, −16 mm, −6 mm, and 4 mm, respectively. The bottom row shows the results from six different sagittal slices, where the x-coordinates in MNI space from left to right are x = −35 mm, −20 mm, −5 mm, 10 mm, 25 mm, and 40 mm, respectively. Red-yellow clusters show the significant positive correlation of FA with MCS for all participants in the GO-ME/CFS group.

**Supplementary Fig. 4.**
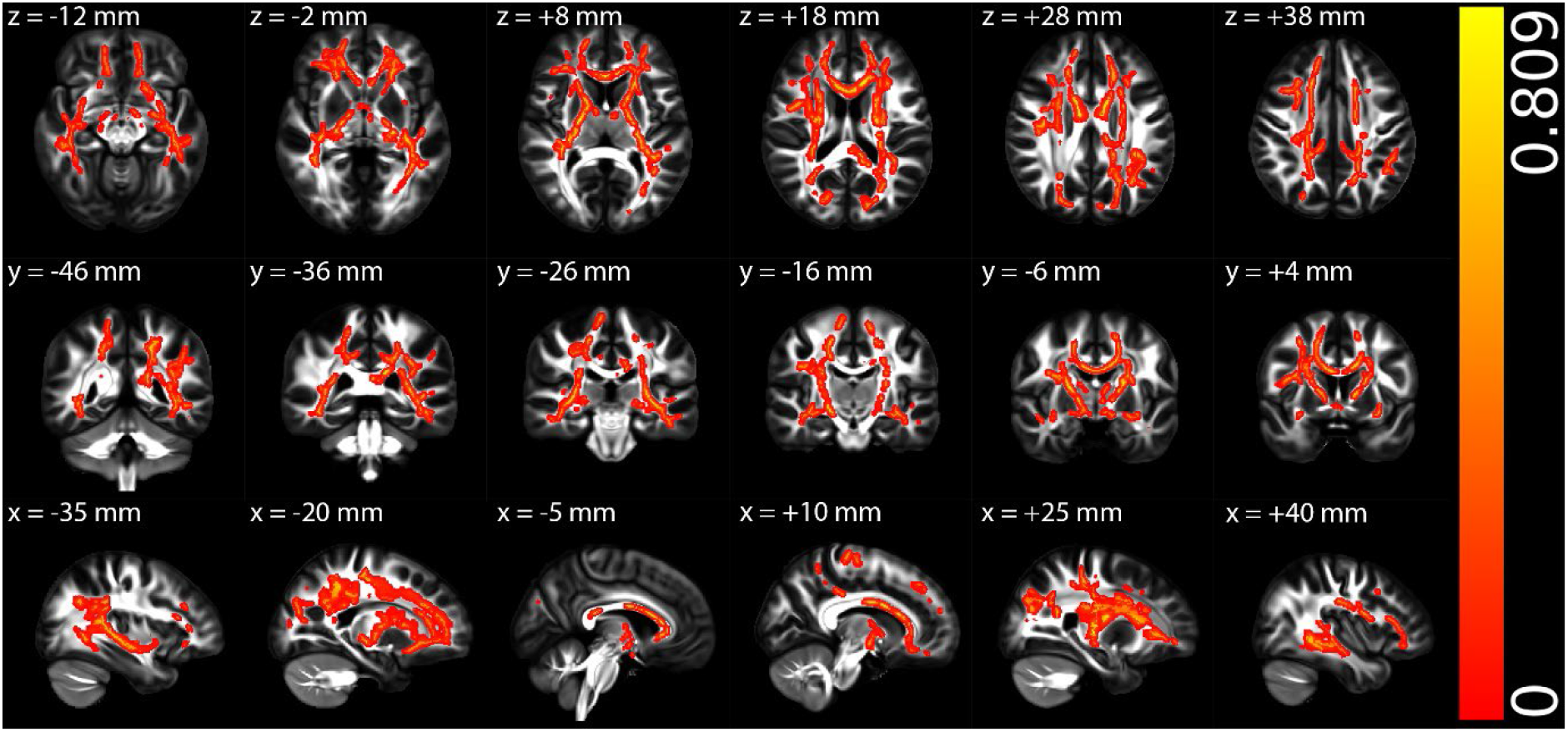
The multiple regression results of radial diffusivity (RD) with disease severity for gradual onset ME/CFS (GO-ME/CFS) patient group in Montreal Neurological Institute (MNI) 152 standard space based on the reference FSL_HCP1065 FA 1×1x1mm standard-space image. The top row shows the results from six different axial slices, where the z-coordinates in MNI space from left to right are z = −12 mm, −2 mm, 8 mm, 18 mm, 28 mm, and 38 mm, respectively. The middle row shows the results from six different coronal slices, where the y-coordinates in MNI space from left to right are y = −46 mm, −36 mm, −26 mm, −16 mm, −6 mm, and 4 mm, respectively. The bottom row shows the results from six different sagittal slices, where the x-coordinates in MNI space from left to right are x = −35 mm, −20 mm, −5 mm, 10 mm, 25 mm, and 40 mm, respectively. Red-yellow clusters show the significant positive correlation of RD with disease severity for GO-ME/CFS patient group.

**Supplementary Fig. 5.**
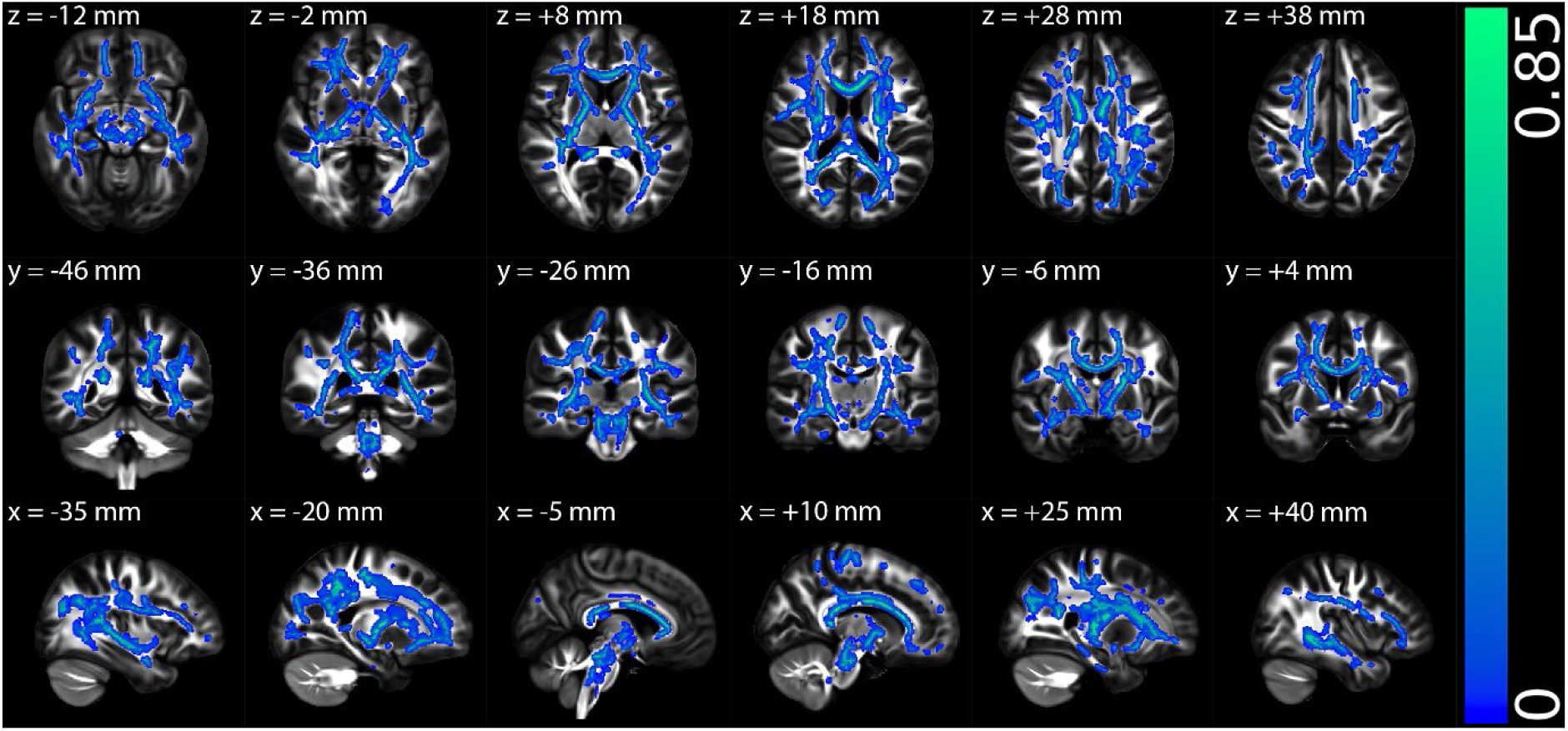
The multiple regression results of fractional anisotropy (FA) with disease severity for gradual onset ME/CFS (GO-ME/CFS) patient group in Montreal Neurological Institute (MNI) 152 standard space based on the reference FSL_HCP1065 FA 1×1x1mm standard-space image. The top row shows the results from six different axial slices, where the z-coordinates in MNI space from left to right are z = −12 mm, −2 mm, 8 mm, 18 mm, 28 mm, and 38 mm, respectively. The middle row shows the results from six different coronal slices, where the y-coordinates in MNI space from left to right are y = −46 mm, −36 mm, −26 mm, −16 mm, −6 mm, and 4 mm, respectively. The bottom row shows the results from six different sagittal slices, where the x-coordinates in MNI space from left to right are x = −35 mm, −20 mm, −5 mm, 10 mm, 25 mm, and 40 mm, respectively. Blue-green clusters show the significant negative correlation of FA with disease severity for GO-ME/CFS patient group.

**Supplementary Fig. 6.**
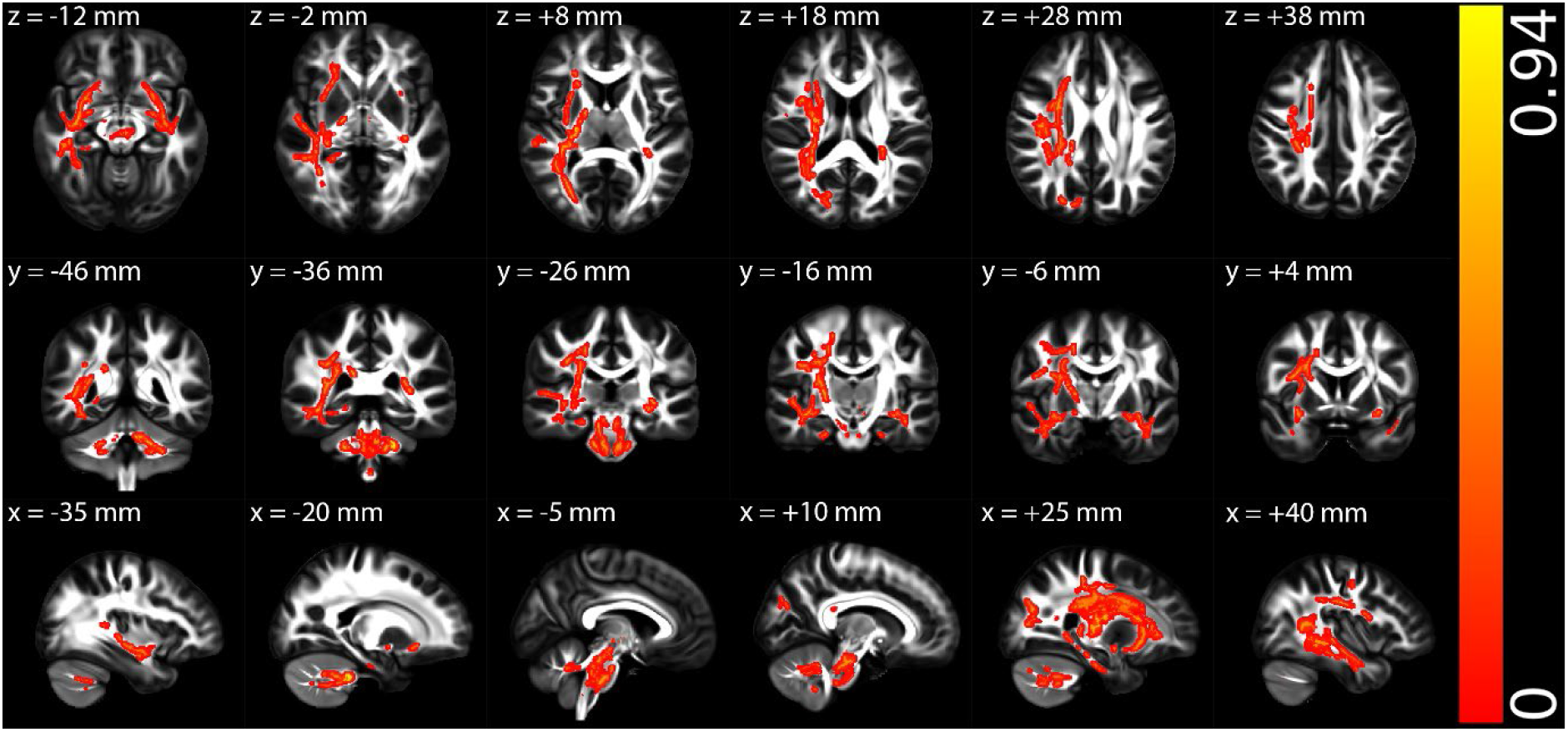
The tract-based spatial statistics (TBSS) results of axial diffusivity (AD) between the healthy control and post-infectious ME/CFS (PI-ME/CFS) patient groups in Montreal Neurological Institute (MNI) 152 standard space based on the reference FSL_HCP1065 fractional anisotropy (FA) 1×1x1mm standard-space image. The top row shows the results from six different axial slices, where the z-coordinates in MNI space from left to right are z = −12 mm, −2 mm, 8 mm, 18 mm, 28 mm, and 38 mm, respectively. The middle row shows the results from six different coronal slices, where the y-coordinates in MNI space from left to right are y = −46 mm, −36 mm, −26 mm, −16 mm, −6 mm, and 4 mm, respectively. The bottom row shows the results from six different sagittal slices, where the x-coordinates in MNI space from left to right are x = −35 mm, −20 mm, −5 mm, 10 mm, 25 mm, and 40 mm, respectively. Red-yellow clusters show the significant increased AD in PI-ME/CFS patients.

**Supplementary Fig. 7.**
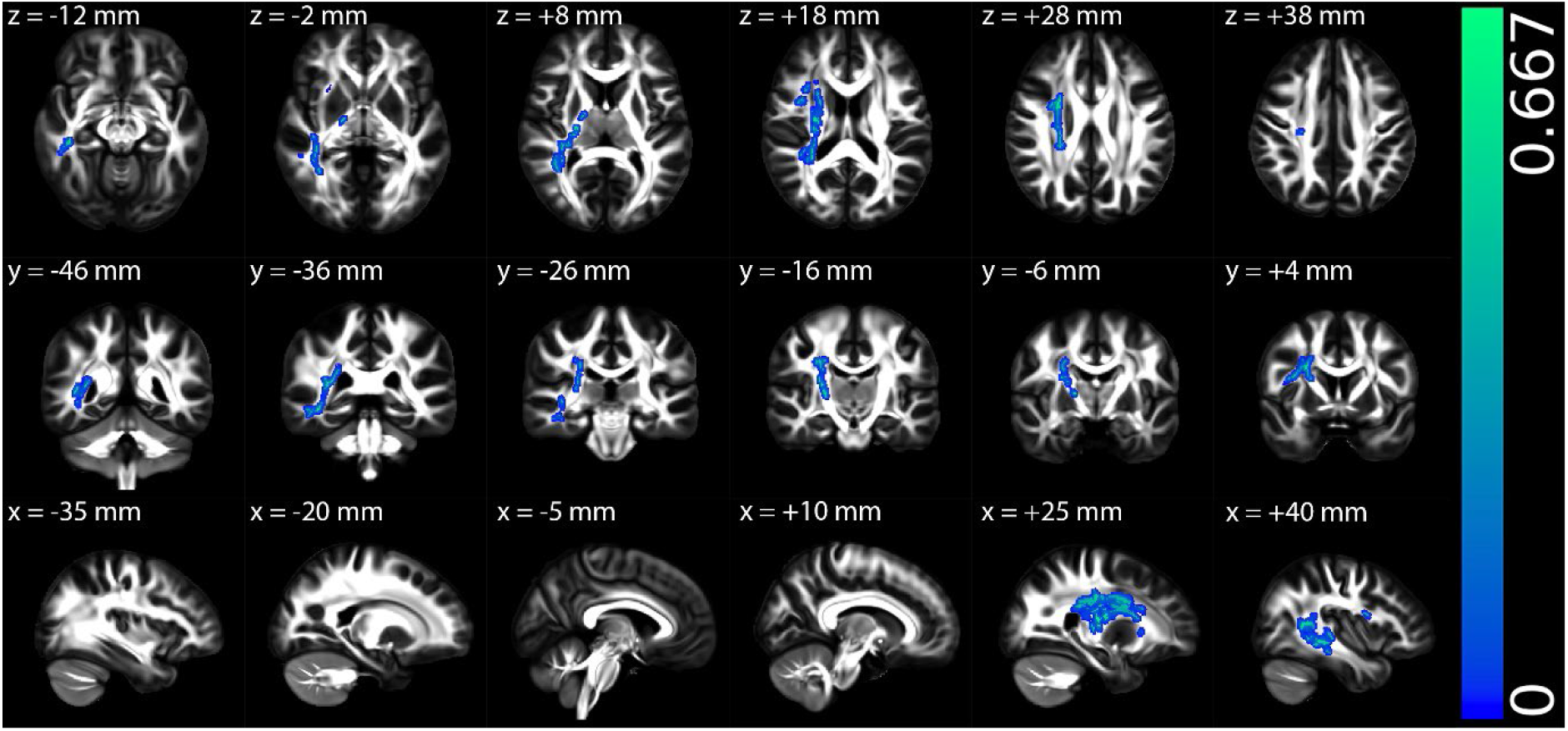
The multiple regression results of axial diffusivity (AD) with physical component summary (PCS) among all eligible participants in the post-infectious ME/CFS (PI- ME/CFS) group in Montreal Neurological Institute (MNI) 152 standard space based on the reference FSL_HCP1065 fractional anisotropy (FA) 1×1x1mm standard-space image. The top row shows the results from six different axial slices, where the z-coordinates in MNI space from left to right are z = −12 mm, −2 mm, 8 mm, 18 mm, 28 mm, and 38 mm, respectively. The middle row shows the results from six different coronal slices, where the y-coordinates in MNI space from left to right are y = −46 mm, −36 mm, −26 mm, −16 mm, −6 mm, and 4 mm, respectively. The bottom row shows the results from six different sagittal slices, where the x-coordinates in MNI space from left to right are x = −35 mm, −20 mm, −5 mm, 10 mm, 25 mm, and 40 mm, respectively. Blue-green clusters show the significant negative correlation of AD with PCS for all eligible participants in the PI-ME/CFS group.

**Supplementary Fig. 8.**
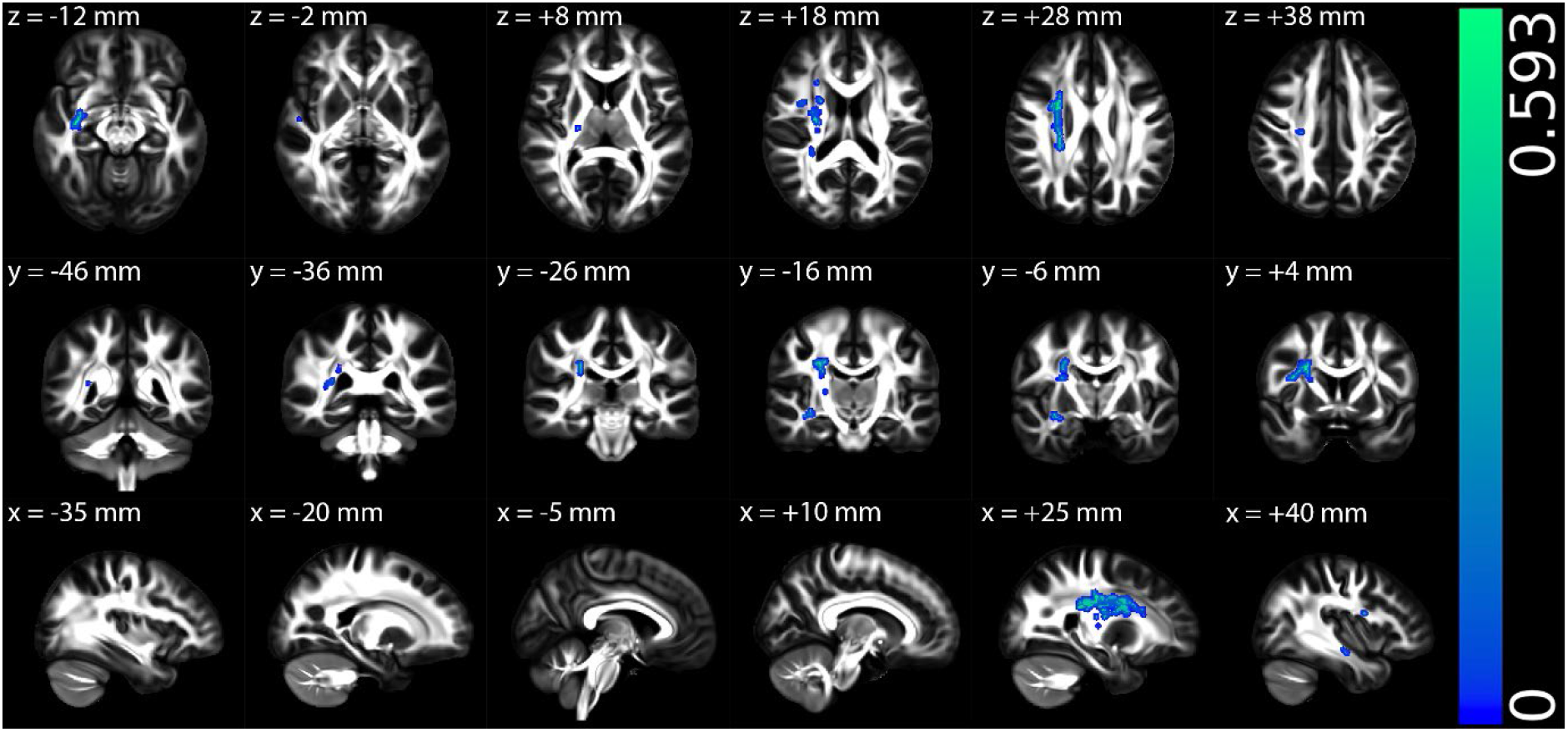
The multiple regression results of axial diffusivity (AD) with Bell’s Disability Scale (BDS) among all eligible participants in the post-infectious ME/CFS (PI- ME/CFS) group in Montreal Neurological Institute (MNI) 152 standard space based on the reference FSL_HCP1065 fractional anisotropy (FA) 1×1x1mm standard-space image. The top row shows the results from six different axial slices, where the z-coordinates in MNI space from left to right are z = −12 mm, −2 mm, 8 mm, 18 mm, 28 mm, and 38 mm, respectively. The middle row shows the results from six different coronal slices, where the y-coordinates in MNI space from left to right are y = −46 mm, −36 mm, −26 mm, −16 mm, −6 mm, and 4 mm, respectively. The bottom row shows the results from six different sagittal slices, where the x- coordinates in MNI space from left to right are x = −35 mm, −20 mm, −5 mm, 10 mm, 25 mm, and 40 mm, respectively. Blue-green clusters show the significant negative correlation of AD with BDS for all eligible participants in the PI-ME/CFS group.

**Supplementary Fig. 9.**
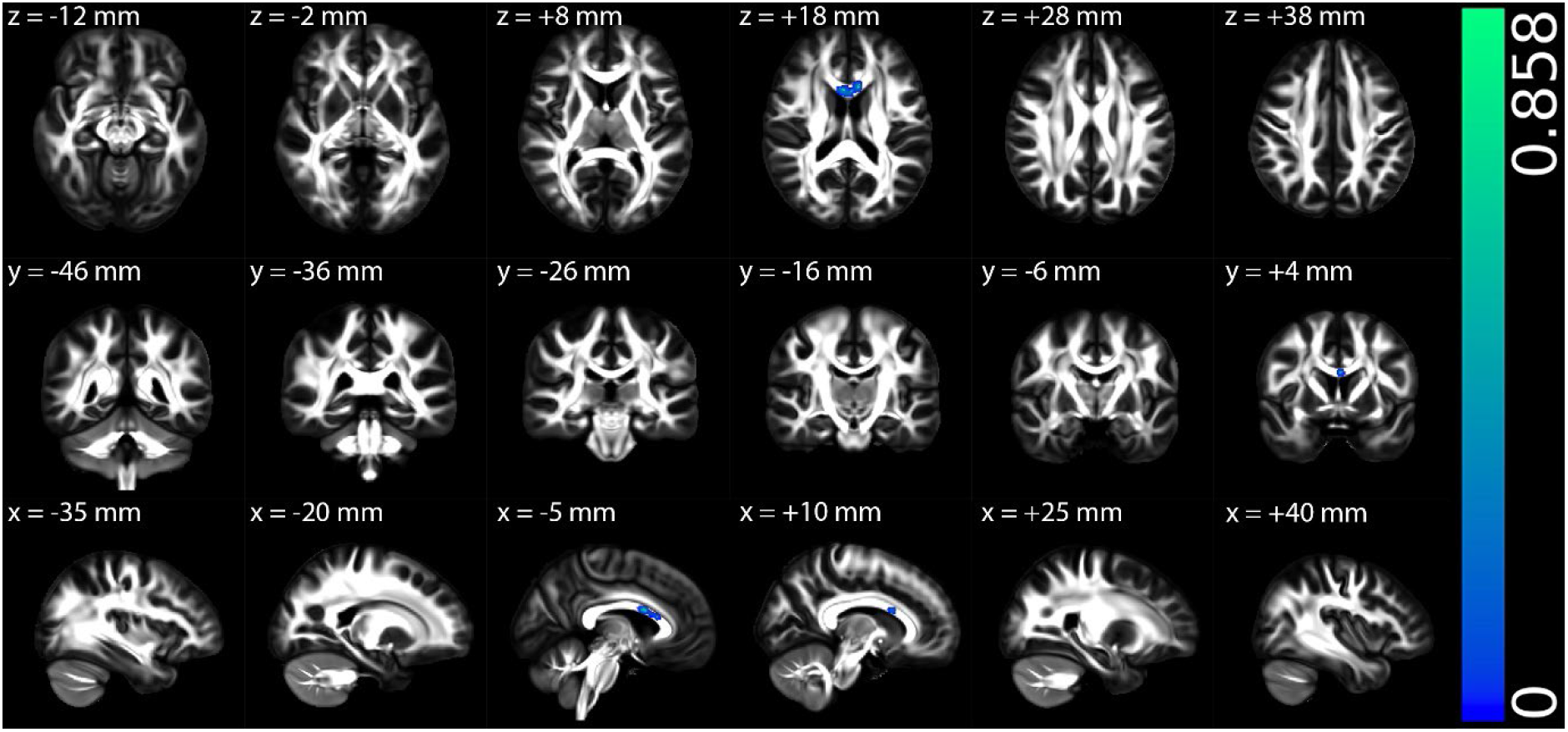
The tract-based spatial statistics (TBSS) results of axial diffusivity (AD) between the healthy control and gradual onset ME/CFS (GO-ME/CFS) patient groups in Montreal Neurological Institute (MNI) 152 standard space based on the reference FSL_HCP1065 fractional anisotropy (FA) 1×1x1mm standard-space image. The top row shows the results from six different axial slices, where the z-coordinates in MNI space from left to right are z = −12 mm, −2 mm, 8 mm, 18 mm, 28 mm, and 38 mm, respectively. The middle row shows the results from six different coronal slices, where the y-coordinates in MNI space from left to right are y = −46 mm, −36 mm, −26 mm, −16 mm, −6 mm, and 4 mm, respectively. The bottom row shows the results from six different sagittal slices, where the x-coordinates in MNI space from left to right are x = −35 mm, −20 mm, −5 mm, 10 mm, 25 mm, and 40 mm, respectively. Blue-green clusters show the significant decreased AD in GO-ME/CFS patients.

**Supplementary Fig. 10.**
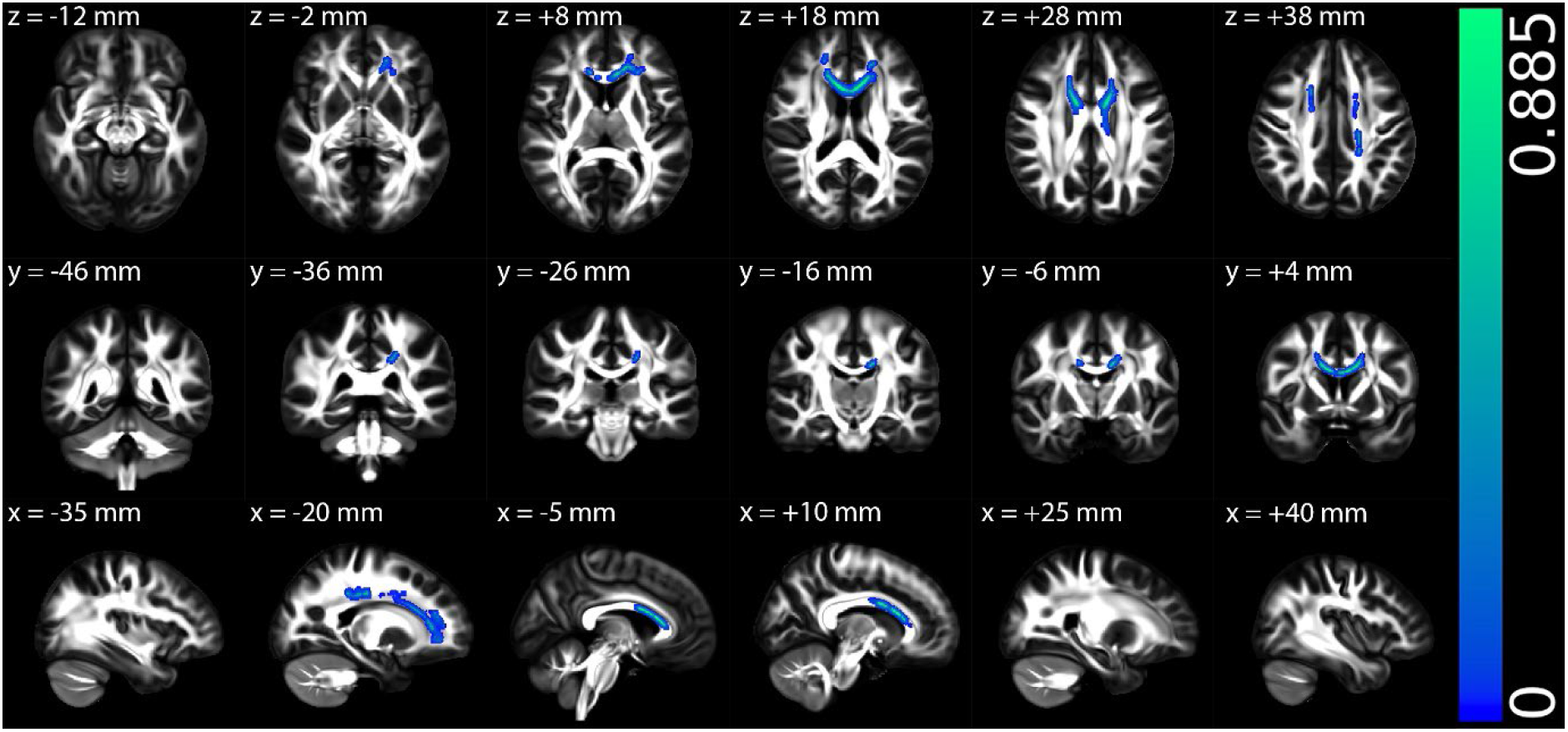
The tract-based spatial statistics (TBSS) results of mean diffusivity (MD) between the healthy control and gradual onset ME/CFS (GO-ME/CFS) patient groups in Montreal Neurological Institute (MNI) 152 standard space based on the reference FSL_HCP1065 fractional anisotropy (FA) 1×1x1mm standard-space image. The top row shows the results from six different axial slices, where the z-coordinates in MNI space from left to right are z = −12 mm, −2 mm, 8 mm, 18 mm, 28 mm, and 38 mm, respectively. The middle row shows the results from six different coronal slices, where the y-coordinates in MNI space from left to right are y = −46 mm, −36 mm, −26 mm, −16 mm, −6 mm, and 4 mm, respectively. The bottom row shows the results from six different sagittal slices, where the x-coordinates in MNI space from left to right are x = −35 mm, −20 mm, −5 mm, 10 mm, 25 mm, and 40 mm, respectively. Blue-green clusters show the significant decreased MD in GO-ME/CFS patients.

**Supplementary Fig. 11.**
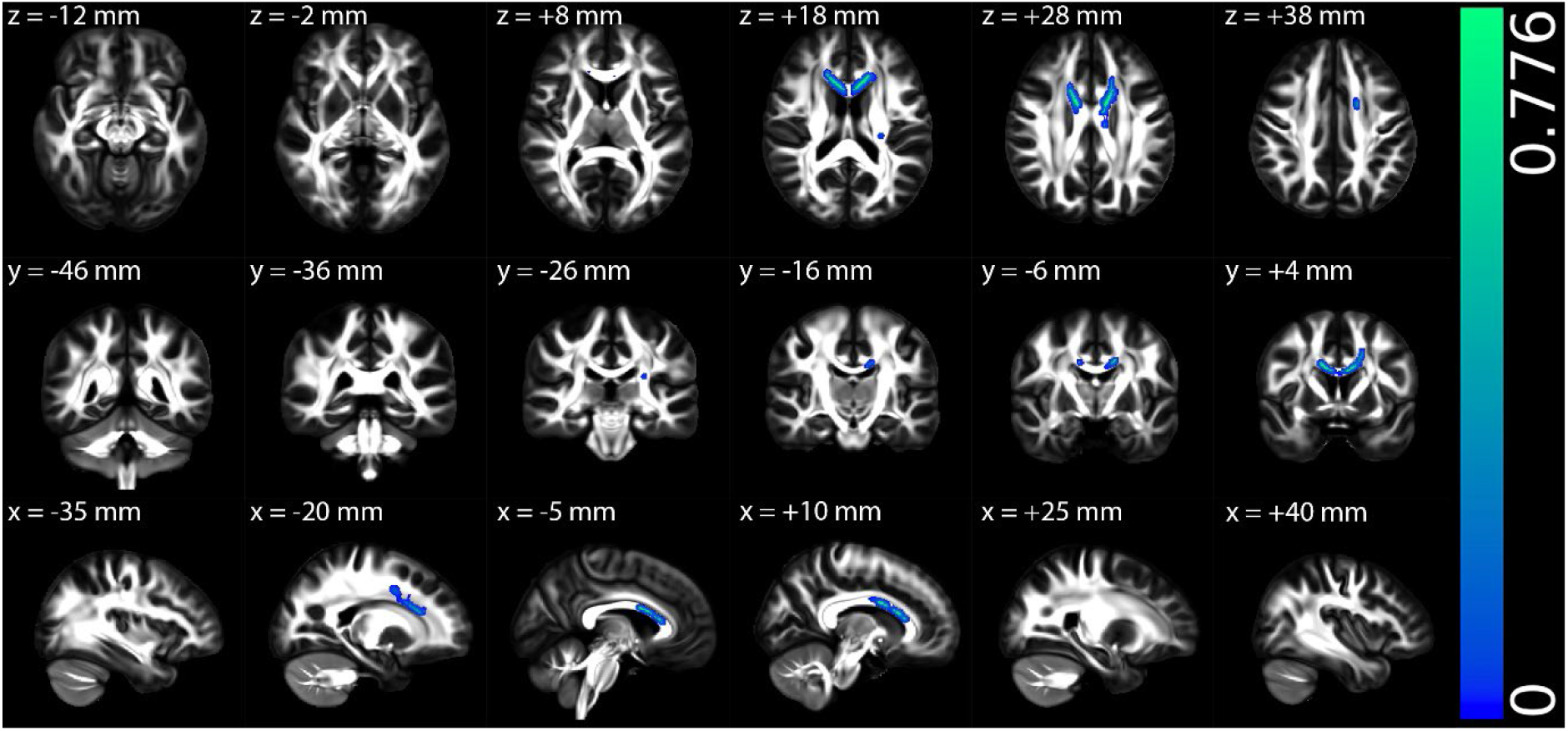
The tract-based spatial statistics (TBSS) results of radial diffusivity (RD) between the healthy control and gradual onset ME/CFS (GO-ME/CFS) patient groups in Montreal Neurological Institute (MNI) 152 standard space based on the reference FSL_HCP1065 fractional anisotropy (FA) 1×1x1mm standard-space image. The top row shows the results from six different axial slices, where the z-coordinates in MNI space from left to right are z = −12 mm, −2 mm, 8 mm, 18 mm, 28 mm, and 38 mm, respectively. The middle row shows the results from six different coronal slices, where the y-coordinates in MNI space from left to right are y = −46 mm, −36 mm, −26 mm, −16 mm, −6 mm, and 4 mm, respectively. The bottom row shows the results from six different sagittal slices, where the x-coordinates in MNI space from left to right are x = −35 mm, −20 mm, −5 mm, 10 mm, 25 mm, and 40 mm, respectively. Blue-green clusters show the significant decreased RD in GO-ME/CFS patients.

**Supplementary Table 1:** Demographic and behavioural information of all eligible participants in the post-infectious ME/CFS (PI-ME/CFS) group before age, sex, MET and number matching (1:1)

| Characteristics | Patients with PI-ME/CFS | Healthy controls | P Values |
| --- | --- | --- | --- |
| **N** | 43 | 58 | n/a |
| **Female/male** | 37/6 | 50/8 | n/a |
| **Age (years)** | 42.58 ± 12.41 [24, 65] | 37.22 ± 11.03 [19, 62] | 0.07 |
| **BMI (kg/m<sup>2</sup>)** | 25.16 ± 4.16 [17.18, 34.66] | 24.74 ± 4.22 [17.36, 35.38] | 0.48 |
| **MET rate** | 1.29 ± 0.14 [1.04, 1.74] | 1.32 ± 0.14 [1, 1.74] | 0.07 |
| **HADS-anxiety** | 7.40 ± 4.01 [0, 15] | 4.59 ± 3.13 [0, 16] | < 0.001 |
| **HADS-depression** | 6.21 ± 3.80 [0, 16] | 2.02 ± 2.76 [0, 11] | < 0.001 |
| **SF-36 - MCS** | 44.52 ± 7.03 [30.29, 58.09] | 53.28 ± 6.00 [37.55, 62.46] | < 0.001 |
| **SF-36 - PCS** | 35.87 ± 9.98 [15.83, 57.23] | 58.43 ± 6.82 [35.43, 68.30] | < 0.001 |
| **Global PSQI** | 9.15 ± 3.32 [2, 17] | 4.78 ± 3.14 [0, 14] | < 0.001 |
| **BDS** | 42.56 ± 15.44 [20, 90] | 94.83 ± 12.60 [40, 100] | < 0.001 |
| **Disease severity** | 3.12 ± 1.26 [1, 5] | n/a | n/a |

Note – The data are presented as mean ± standard [lower bound, higher bound]. ME/CFS = myalgic encephalomyelitis/chronic fatigue syndrome; BMI = body mass index; MET = metabolic equivalents (one MET is defined as the energy used in resting or sitting still); HADS = Hospital Anxiety and Depression Scale; SF-36 = 36-item Short-Form; MCS = mental component summary; PCS = physical component summary; PSQI = Pittsburgh Sleep Quality Index; BDS = Bell's Disability Scale. Disease severity score 1 is referred to as Mild, score 2 is referred to as Mild-Moderate, score 3 is referred to as Moderate, score 4 is referred to as Moderate-Severe, and score 5 is referred to as Severe.

**Supplementary Table 2:** Demographic and behavioural information of all eligible participants in the gradual onset ME/CFS (GO-ME/CFS) group before age, sex, MET and number matching (1:1)

| Characteristics | Patients with GO-ME/CFS | Healthy controls | P Values |
| --- | --- | --- | --- |
| **N** | 33 | 66 | n/a |
| **Female/male** | 26/7 | 52/14 | n/a |
| **Age (years)** | 42.03 ± 12.87 [24, 62] | 37.09 ± 11.14 [19, 62] | 0.10 |
| **BMI (kg/m<sup>2</sup>)** | 24.98 ± 4.75 [18.69, 34.89] | 25.08 ± 4.26 [17.36, 35.38] | 0.86 |
| **MET rate** | 1.30 ± 0.15 [1.04, 1.72] | 1.35 ± 0.15 [1, 1.74] | 0.07 |
| **HADS-anxiety** | 8.97 ± 5.21 [1, 18] | 4.64 ± 3.20 [0, 16] | < 0.001 |
| **HADS-depression** | 7.21 ± 4.41 [1, 17] | 2.08 ± 2.72 [0, 11] | < 0.001 |
| **SF-36 - MCS** | 42.95 ± 9.03 [23.34, 56.77] | 53.26 ± 6.14 [37.18, 62.46] | < 0.001 |
| **SF-36 - PCS** | 37.56 ± 6.49 [27.10, 50.68] | 58.58 ± 6.55 [35.43, 68.30] | < 0.001 |
| **Global PSQI** | 9.09 ± 3.39 [3, 15] | 4.58 ± 3.04 [0, 14] | < 0.001 |
| **BDS** | 43.94 ± 14.35 [10, 70] | 95.00 ± 12.06 [40, 100] | < 0.001 |
| **Disease severity** | 2.76 ± 1.06 [1, 4] | n/a | n/a |

Note – The data are presented as mean ± standard [lower bound, higher bound]. ME/CFS = myalgic encephalomyelitis/chronic fatigue syndrome; BMI = body mass index; MET = metabolic equivalents (one MET is defined as the energy used in resting or sitting still); HADS = Hospital Anxiety and Depression Scale; SF-36 = 36-item Short-Form; MCS = mental component summary; PCS = physical component summary; PSQI = Pittsburgh Sleep Quality Index; BDS = Bell's Disability Scale. Disease severity score 1 is referred to as Mild, score 2 is referred to as Mild-Moderate, score 3 is referred to as Moderate, score 4 is referred to as Moderate-Severe, and score 5 is referred to as Severe.

